# Phenotypes associated with genetic determinants of type I interferon regulation in the UK Biobank: a protocol

**DOI:** 10.1101/2023.10.12.23296935

**Authors:** Bastien Rioux, Michael Chong, Rosie Walker, Sarah McGlasson, Kristiina Rannikmäe, Daniel McCartney, John McCabe, Robin Brown, Yanick J Crow, David Hunt, William Whiteley

## Abstract

**Introduction:** Type I interferons are cytokines involved in innate immunity against viruses. Genetic disorders of type I interferon regulation are associated with a range of autoimmune and cerebrovascular phenotypes. Carriers of pathogenic variants involved in genetic disorders of type I interferons are generally considered asymptomatic. Preliminary data suggests, however, that genetically determined dysregulation of type I interferon responses is associated with autoimmunity, and may also be relevant to sporadic cerebrovascular disease and dementia. We aim to determine whether functional variants in genes involved in type I interferon regulation and signalling are associated with the risk of autoimmunity, stroke, and dementia in a population cohort.

**Methods and analysis:** We will perform a hypothesis-driven candidate pathway association study of type I interferon-related genes using rare variants in the UK Biobank (UKB). We will manually curate type I interferon regulation and signalling genes from a literature review and Gene Ontology, followed by clinical and functional filtering. Variants of interest will be included based on pre-defined clinical relevance and functional annotations (using LOFTEE, M-CAP and a minor allele frequency <0.1%). The association of variants with 15 clinical and three neuroradiological phenotypes will be assessed with a rare variant genetic risk score and gene-level tests, using a Bonferroni-corrected p-value threshold from the number of genetic units and phenotypes tested. We will explore the association of significant genetic units with 196 additional health-related outcomes to help interpret their relevance and explore the clinical spectrum of genetic perturbations of type I interferon.

**Ethics and dissemination:** The UKB has received ethical approval from the North West Multicentre Research Ethics Committee, and all participants provided written informed consent at recruitment. This research will be conducted using the UKB Resource under application number 93,160. We expect to disseminate our results in a peer-reviewed journal and at an international cardiovascular conference.

**STRENGTHS AND LIMITATIONS OF THIS STUDY:**

- The UK Biobank is the largest whole-exome sequencing project to date, with marked power to detect associations from a limited number of rare, functional variants.
- Our study will leverage current knowledge of interferon biology and genotype-phenotype correlations in Mendelian diseases of type I interferon to test biologically plausible hypotheses.
- The UK Biobank includes phenotypes from multiple sources, which improves classification accuracy for several health outcomes such as stroke and dementia.
- We will carefully select genes and variants with strong evidence of biological relevance to optimize the power of our analyses, which is particularly relevant for less common phenotypes in the UK Biobank such as systemic lupus erythematosus.
- We will increase the specificity of predicted loss-of-function variants by using stringent sample quality control and filtering criteria.

## INTRODUCTION

Interferons are a family of innate inflammatory cytokines primarily secreted by host cells in response to viruses (type I: mainly interferon-α and -β; type II: interferon-γ; type III: interferon-λ). Interferon-stimulated genes are involved in a wide range of processes, namely cellular defence against pathogens, apoptosis, nucleic acid degradation, and cell-to-cell communication [1]. Defects in type I interferon homeostasis are associated with autoimmunity, being implicated in the pathogenesis of systemic lupus erythematosus and other autoimmune disorders such as rheumatoid arthritis, Sjögren’s syndrome, and scleroderma [2]. Low-grade type I interferon upregulation may also contribute to sporadic cerebrovascular disease and dementia. Preclinical data suggest type I interferon-related vascular inflammation is an essential contributor to atherosclerosis and may be involved in cerebral small vessel disease [3, 4]. Stroke risk is increased after long-term exposure to exogenous recombinant type I interferon [4, 5], whereas white matter hyperintensities (a radiological manifestation of cerebral small vessel disease), large vessel disease and stroke are more frequent in people with systemic lupus erythematosus as compared to the general population [6, 7].

Genetic type I interferonopathies are a group of rare Mendelian autoinflammatory diseases hypothesised to be caused by an upregulation of type I interferons. Affected individuals with Aicardi-Goutières syndrome, the first type I interferonopathy described, most frequently present in early childhood with progressive encephalopathy, skin vasculopathy, and autoimmunity [8], in addition to prominent white matter hyperintensities, calcifications and large vessel disease (aneurysms, arterial calcifications, stenoses) on brain imaging [9]. Most, albeit not all (e.g., mutations in *IFIH1*, *STING* and *COPA*), pathogenic variants associated with type I interferonopathies result in a loss-of-function (LOF) of key interferon negative regulators inherited as autosomal recessive traits. Carriers of such pathogenic variants are generally considered asymptomatic, although growing evidence from case series suggests they may also exhibit high expression of interferon-stimulated genes [10] and have mild interferonopathy-related traits [11, 12]. Uncertainty remains, however, as to whether carriers of pathogenic variants in genes involved in type I interferon signalling and regulation have an increased risk of interferonopathy-related phenotypes such as autoimmunity, cerebrovascular disease, and dementia. Moreover, the causal role of type I interferon in sporadic cerebrovascular disease and dementia has not been comprehensively assessed in a population-based study [13], and whether findings from preclinical studies and observations in conditions with impaired interferon homeostasis translate to the general population is unclear.

We will apply a candidate pathway approach to determine whether functional variants in genes involved in type I interferon regulation and signalling are associated with clinical and neuroradiological interferonopathy phenotypes in the general population. We hypothesize that a subset of rare functional variants that result in an upregulation of the type I interferon cascade are associated with core interferonopathy phenotypes.

## METHODS AND ANALYSIS

We will report our results using guidance from the Strengthening the Reporting of Genetic Association Studies (STREGA) initiative [14], and present the protocol checklist in **Supplemental methods 1**. We present a graphical abstract of our protocol in **Figure 1**.

**Figure 1.**
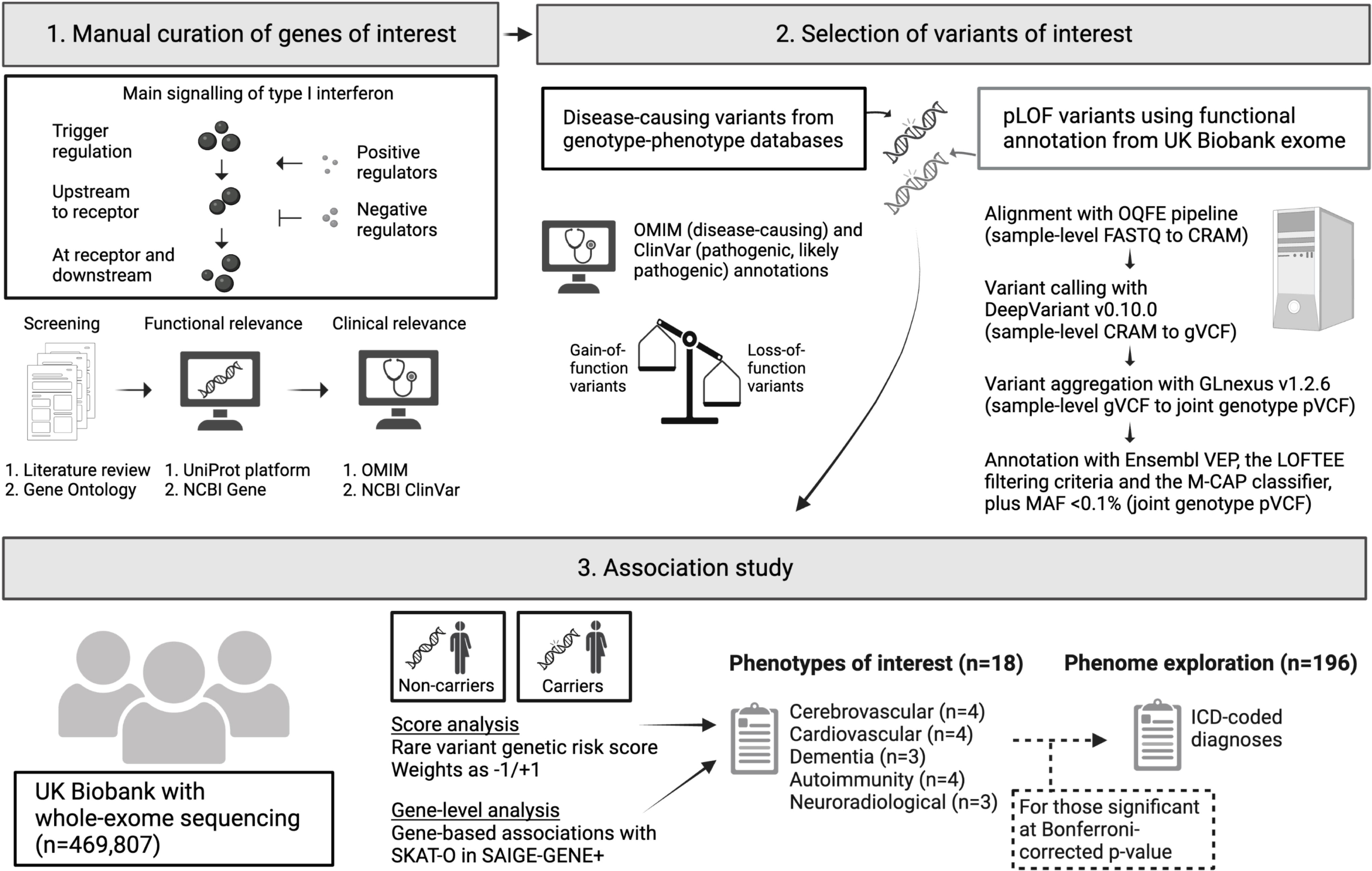
Graphical summary of the study methodology. This summary illustrates the three main steps of the study: i) genes of interest will be identified from a literature review and Gene Ontology, followed by clinical and functional filtering, ii) variants of interest will be included based on their clinical relevance and functional annotations, and iii) the association of variants and phenotypes will be tested with a rare variant genetic risk score and gene-level tests. Abbreviations: LOFTEE, Loss-Of-Function Transcript Effect Estimator; M-CAP, Mendelian Clinically Applicable Pathogenicity score; NCBI, National Center for Biotechnology Information; OMIM, Online Mendelian Inheritance in Man; OQFE, Original Quality Functionally Equivalent; pLOF, predicted loss-of-function; SKAT-O, optimal sequence kernel association test; VEP, Variant Effect Predictor. Created with BioRender.com.

### Study population and exome extraction

We will use data from the UK Biobank (UKB), a large population-based cohort of 502,650 participants mostly of white British ancestry who were aged 40-69 years when recruited from UK patient registries between 2006 and 2010 (response rate: 5.5%) [15, 16]. We will consider individuals with whole-exome sequencing based on the final exome data release (July 2022; n=469,807; 93.5% of participants). The exome was sequenced in two batches composed of the first 50k participants (phase 1) and all other samples (phase 2). Participants in the first phase were selected to enrich certain phenotypes, which may lead to spurious associations given time-varying sequencing coverage if this batch effect is not controlled (see below).

Genomic DNA samples were sent to the Regeneron Genetics Centre (Tarrytown, New York, USA) as part of a collaboration with the UKB and stored at -80°C. Genomic libraries with a mean fragment size of 200 base pairs (bp) were created enzymatically and tagged with barcodes of 10 bp before capture. Exome was obtained by next generation sequencing using the Illumina NovaSeq 6000 platform (S2 and S4 flow cells for the first and second phase, respectively) and a target-enrichment probe kit (IDT xGen® Exome Research Panel v1.0) to enable deep and uniform coverage of ∼39 Mbp (19,396 genes).

### Whole-exome sequencing data

We will use the multi-sample project-level VCF (pVCF) files made available by the UKB [17]. To obtain these joint genotype data, raw sequencing outputs (FASTQs) were initially processed into sample-level aligned sequences (CRAMs) with a standard protocol (the Original Quality Functionally Equivalent; OQFE), which maps short sequences to the GRCh38 reference genome with alternate loci and marks duplicate segments [18]. DeepVariant (v0.10.0) was used to call variants from sample-level CRAMs and produce variant call data (gVCF) for each participant [19]. This calling approach uses a deep convolutional neural network to determine the most likely genotype at each locus from the reference genome, base reads and quality scores [20]. It outperforms existing state-of-the-art tools to call single nucleotide variants (SNVs) and small insertions or deletions (indels; up to 50 bp by definition), achieving high overall accuracy (>99.5%) [20, 21]. The variant call data set includes exome capture targets and their immediate flanking regions (100 bp upstream and downstream of each target). Sample-level variants were aggregated into joint genotype pVCF files with a standard analysis pipeline (GLnexus v1.2.6) [19, 22].

For quality control, we will exclude participants with a mismatch between their genetically recorded and self-reported sex or with sex chromosome aneuploidy (∼0.2%). We will apply a set of per-variant quality control metrics as previously employed for the UKB exome to analyse variants with [23]:

i. individual and variant missingness <10%;
ii. Hardy Weinberg equilibrium p-value >10^-15^;
iii. at least one sample per site with allele balance threshold >0.15 for SNVs and >0.20 for small indels;
iv. minimum read coverage depth of seven for SNVs and 10 for indels.

We will also use a sequencing depth ≥10x in 90% of samples for our rare variant analysis, to prevent spurious associations that may result from batch effect [24]. We will resolve haplotype phase with the Segmented HAPlotype Estimation and Imputation Tools version 5 (SHAPEIT5 v1.0.0), which phases rare variants from the UKB with high accuracy (switch error rate <5% with minor allele count >5) [25].

### Genes of interest

We will apply a hypothesis-driven candidate pathway approach of type I interferon-related genes by adapting a previously described methodology [26]. We will consider for inclusion any gene encoding a protein of interest belonging to one of the three following categories:

1. A negative regulator, positive regulator, or effector along the main signalling pathway of type I interferon (**Figure 2**);
2. A protein directly affecting the activity of an interferon regulator or effector (e.g., E3 ubiquitin-protein ligase TRIM21 inhibits interferon regulatory factor 3, a transcription factor that controls multiple type I interferon-inducing pathways; both proteins are therefore considered for inclusion);
3. A protein involved in genetic type I interferonopathies (see **Supplemental table 1**).

**Figure 2.**
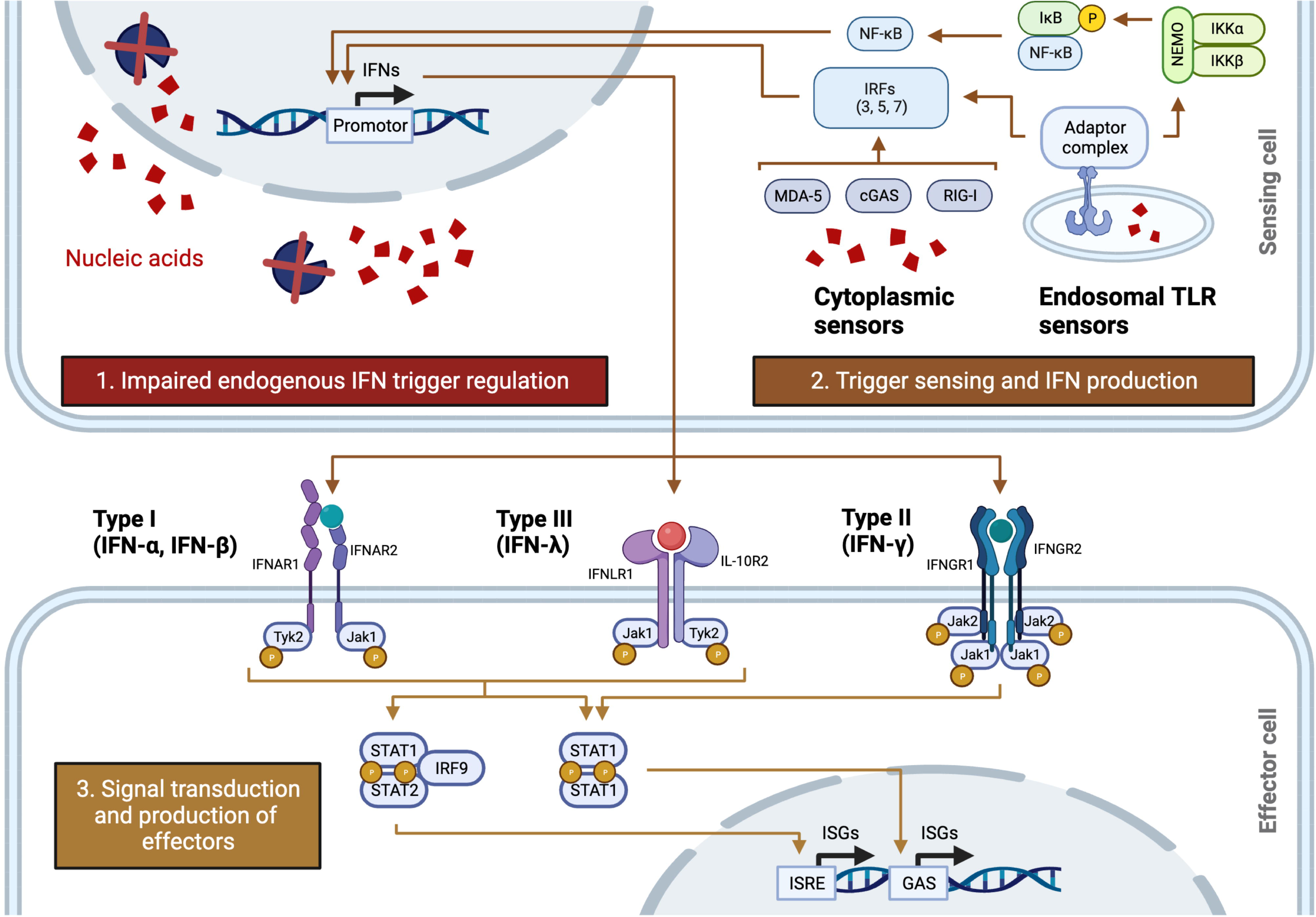
Overview of the interferon cascade. Graphical overview of the main steps involved in interferon regulation and signalling. Endogenous nucleases (blue circle sectors) remove nucleic acids (red confetti) that can trigger interferon production. Abnormal accumulation of endogenous material through impaired regulation (box 1) and viral nucleic acids (not shown) can trigger interferon production through linkage to i) toll-like receptor sensors at the cell membrane surface (not shown) and at endosomes, and ii) cytoplasmic sensors (box 2). Interferons are sensed by cell surface receptors specific to types I (heterodimer with subunits IFNAR1 and IFNAR2), II (heterotetramer with two IFNGR1 and two IFNGR2 subunits) and III (heterodimer with subunits IFNLR1 and IL-10R2) ligands. Signal transduction and intracellular signalling through JAK-STAT activates the transcription of interferon-stimulated genes (box 3). Abbreviations: GAS, gamma-activated sequence; IFN, interferon; IRF, interferon regulatory factor; ISG, interferon-stimulated gene; ISRE, interferon-stimulated response element; TLR, toll-like receptor. Created with BioRender.com.

We did not consider regulatory proteins acting beyond the second order of regulation (e.g., regulators of E3 ubiquitin-protein ligase TRIM21) to adequately balance the need to include important regulators of type I interferon, while maintaining their specificity to type I interferon signalling.

We will produce a preliminary list of genes from i) recently published reviews on type I interferon biology and ii) annotations in Gene Ontology [27]. We present herein both completed and upcoming steps. First, we searched Ovid MEDLINE to identify reviews describing ≥2 proteins of interest in physiological conditions. We used interferons (of any type to increase the sensitivity of our search) and regulation as main concepts, in addition to a previously published hedge for reviews (**Table 1**) [28]. We queried MEDLINE from January 2000 onwards to only include recent reviews, and conducted our search in English as we expected reviews in other languages to present similar information. Our strategy yielded 194 records. A single author (BR) will screen records by title and abstract, and include relevant articles after full-text reading. We will manually add four recent reviews [8, 9, 29, 30] on genetic interferonopathies to ensure these genes are captured. A single author (BR) will extract relevant proteins, their corresponding genes, and their presumed functions.

**Table 1.** Ovid MEDLINE search strategy.

| Line | Entry | Records |
| --- | --- | --- |
| Interferon concept |  |  |
| 1 | (cytokine* adj (inflammat* or proinflammat*)).tw. | 422 |
| 2 | IFN*.ti. | 15433 |
| 3 | Interferons/ | 25640 |
| 4 | 1 or 2 or 3 | 40607 |
| Regulation concept |  |  |
| 5 | (regulat* or metabolism or biology or function).ti | 1191801 |
| Review design hedge |  |  |
| 6 | meta analysis.mp,pt. or review.pt. or search:.tw. | 3563440 |
| Combine concepts |  |  |
| 7 | 4 and 5 and 6 | 390 |
| 8 | 7 not ((exp animal/ or nonhuman/) not exp human/) | 357 |
| 9 | 8 not (case study/ or case report/) | 356 |
| 10 | limit 9 to dt=20000101-20230110 | 214 |
| 11 | limit 10 to English language | 194 |

This table presents the search strategy conducted in Ovid MEDLINE on 10 January 2023. Abbreviations: adj, adjacent; dt, create date; exp, explode; mp, multi-purpose fields; pt, publication type; ti, text word in title; tw, text word in title and abstract.

Second, we have queried the Gene Ontology resource to validate and enrich our gene set. Gene Ontology provides curated gene-specific knowledge with functional annotation and hierarchical relationships [27, 31, 32]. We extracted a list of 194 genes pertaining to 31 ontology terms relevant to type I interferon (**Supplemental table 2**). We will validate presumed gene product function from reviews and Gene Ontology on the UniProt platform [33] and the National Center for Biotechnology Information (NCBI) Gene database [34] before assigning their function (e.g., negative regulator) and level of action (e.g., downstream to receptors). Discrepancies will be resolved through consensus by three authors with expertise in interferon biology (BR, SM, DH).

From this preliminary list of genes, we will only include those with ≥1 variant associated with a Mendelian disease through any effect on protein function to strengthen their biological relevance. We will search the Online Mendelian Inheritance in Man (OMIM) [35] and the NCBI ClinVar [36] clinical annotation databases for genotype-phenotype correlations. We will validate that all top 21 type I interferon-inducible genes in systemic lupus erythematosus are included in our list, and add missing items [37].

### Variants of interest

We will include both SNVs and small indels in genes of interest with ≥1 of the following protein effects: i) LOF, dominant-negative, or gain-of-function (GOF) disease-causing variants through an autosomal dominant, recessive or X-linked inheritance [38], or ii) predicted LOF variants from functional annotations. We will define disease-causing variants as those reported in ClinVar (as pathogenic or likely pathogenic, excluding variants with conflicting interpretations of pathogenicity), OMIM (as disease-causing), and from discussion with clinical experts in interferonopathies (DH, SM, YC). The protein-level effect will be determined through comments and linked publications in ClinVar, descriptions in OMIM or, if undetermined, inferred from resulting phenotype.

We will also define a second set of putative functional variants identified in UKB participants to increase our statistical power [39]. We will assess the functional impact of these variants on Ensembl with the Variant Effect Predictor (VEP), an online resource that returns annotations on the effect of variants on transcripts and proteins [40]. We will interpret variant pathogenicity with the Loss-Of-Function Transcript Effect Estimator (LOFTEE) and the Mendelian Clinically Applicable Pathogenicity score (M-CAP v1.4). The LOFTEE filtering criteria will be used to annotate non-missense predicted LOF variants, as it provides a conservative filtering strategy to increase specificity (e.g., removal of variants predicted to escape nonsense-mediated decay) and was used to annotate variants in the Genome Aggregation Database (gnomAD; a public resource of ∼126k high-quality exomes from around the world that does not include UKB data) [41] and Genebass (a public resource of exome-based genotype-phenotype associations within the UKB) [42]. The M-CAP score will be used to interpret pathogenicity and nominate missense variants for inclusion [43]. This supervised learning classifier incorporates nine established pathogenicity likelihood scores (namely SIFT and PolyPhen-2) and achieves substantial reduction in the misclassification rate of known pathogenic variants (<5%) as compared to other existing methods (26-38%) [43]. We will define predicted LOF variants as either i) variants that inactivate a protein-coding gene through a premature stop codon, a shift in the transcriptional frame or an alteration of essential splice-site nucleotides (from LOFTEE), or missense variants classified as likely pathogenic (from M-CAP). We will apply a minor allele frequency (MAF) threshold <0.1% in both the UKB and gnomAD to lower the probability of including benign variants and improve our statistical power. Using a more liberal MAF threshold of <1%, 8.03 million SNVs were identified in ∼200k UKB participants, of which 5.4% (∼450k) were predicted LOF variants [23]. In gnomAD, which used LOFTEE without MAF threshold, about 40% of genes had >10 predicted LOF variants [41].

### Phenotypes of interest

We will test the association of selected variants with a set of 15 clinical and three neuroradiological phenotypes of interest in the UKB. These phenotypes were selected based on their frequency in the general population and the UKB, the plausibility of their association with type I interferon upregulation, and from type I interferonopathy clinical presentations (including Mendelian and sporadic diseases). The International Classification of Diseases (ICD) diagnostic codes and UKB fields for each phenotype are presented in **Supplemental table 3**. Genes associated with ≥1 phenotype of interest will be assessed for their association with 196 clinical phenotypes to help interpret their relevance (**Supplemental table 4**). We manually grouped ICD-coded diagnoses by pathophysiology to reduce multiple testing and improve power for less common conditions. As part of the phenome analysis, we will test two stroke definitions developed by Rannikmäe et al [44] to help explain potential misclassifications.

Health-related outcomes were captured through self-completed questionnaires followed by a nurse-led interview on past medical history (at baseline in all and during follow-up for some participants), as well as data linkage with ICD-coded hospital admissions from National Health Service (NHS) registries (primary or secondary diagnoses; ICD v9 and v10) and national death registries (primary and secondary causes of death; ICD v10). Diagnostic codes from primary care (Read codes v2 and v3) are available in a subset of participants (∼45.8%). Cancer diagnoses (ICD v9 and v10) are available through data linkage with national cancer registries. Stroke diagnoses from hospital and death registries have a high sensitivity (point estimate range: 88-94%) and specificity (>99%) [45]. Most stroke cases in the UKB are from hospital and death registries, although ∼27% are self-reported without coded diagnosis [44]. Self-reported strokes have a lower sensitivity (79%) but maintain a high specificity (99%) [46]. In-hospital and death records for all-cause dementia in the UKB have a positive predicted value of ∼85% [47].

We will define phenotypes in the UKB using algorithmically defined (or adjudicated) outcomes (v2.0), first diagnostic occurrences, and cancer registries. Algorithmically defined outcomes are custom diagnostic classification schemes developed by the UKB from self-reports, hospital admissions and death registries to optimize their positive predictive value. First diagnostic occurrences map clinical terms from all available sources into ICD v10 codes (apart from cancer registries). Algorithmically defined outcomes and first diagnostic occurrences will be combined to identify any stroke, ischemic stroke, intracerebral haemorrhage, subarachnoid haemorrhage, all-cause dementia, Alzheimer’s disease, vascular dementia, Parkinson’s disease and myocardial infarction [48]. We chose to combine these two fields to capture primary care events (not included in algorithmically defined outcomes), which is expected to increase the number of cases from 0.7% (n=55) for all-cause dementia to 11.4% (n=1,376) for ischemic stroke. First diagnostic occurrences will be used alone for other non-cancerous conditions. Data from cancer registries will be used to define malignant neoplasms.

We will define three neuroradiological phenotypes: total white matter hyperintensity (WMH) volume, total brain (grey plus white matter) volume, and hippocampal grey matter (mean) volume [49]. Brain magnetic resonance imaging (MRI) scans were obtained in ∼42k participants on 3T Siemens Skyra scanners running VD13A SP4 with a standard Siemens 32-channel radio-frequency receiver head coil. The UKB MRI quality control pipeline includes a pre-processing step to correct for head motion and other artifacts followed by automated identification of equipment failure and excessive artifacts [50]. We will normalize WMH and hippocampal grey matter volumes for head size using the UKB scaling factor derived from the external surface of the skull. The normalized total brain volume is available as an imaging-derived phenotype. We will log-transform WMH volumes given their right-skewed (log-normal) distribution.

The total WMH volume of presumed vascular origin per individual was generated by an image-processing pipeline [50] followed by a segmentation algorithm (the Brain Intensity Abnormality Classification Algorithm tool; BIANCA) using both T1- and T2-weighted/fluid-attenuated inversion recovery (FLAIR) sequences [51]. The algorithm results in high volumetric agreement (intraclass correlation coefficient = 0.99) and very good spatial overlap index (dice similarity index = 0.76) with manual segmentation. Total brain (including the cerebellum and the brainstem, as low as space-based brain masking allows) and regional brain volumes were extracted using tissue-segmented images obtained from an automated algorithm (FMRIB’s Automated Segmentation Tool; FAST) [52] and passed on to the SIENAX analysis pipeline to accurately measure volumetric phenotypes (relative mean error in brain volume = 0.4%) [53]. We will use the mean hippocampal grey matter volume (from right and left hippocampi) as this radiological marker of hippocampal atrophy is associated with memory loss and progression to Alzheimer’s disease [54].

### Statistical analyses

Our primary (score-based) analysis will test the association of all selected genes modelled into a rare variant genetic risk score (RVGRS) with individual phenotypes. We will regress each phenotype on standardized scores using logistic and linear regressions for binary and continuous outcomes, respectively. We will adapt a previously described methodology [55] to define our score as the weighted sum of the number of variants per individual *i* and gene *g* (*V_i,g_*), given a set of *M* genes:

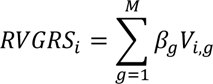

Gene-level weights will be allocated from theoretical variant effects on the type I interferon cascade. For example, LOF variants in genes encoding negative regulators will receive a positive weight (+1) as they are expected to upregulate the cascade, whereas those in genes encoding positive regulators or effectors will receive a negative weight (-1). We chose this conservative weighting method given the technical limitations of weighting variants from a transcriptomic signature (unavailable in the UKB) or a proteomic profile (no measurement of type I interferon in the UKB Olink proteomics).

Our secondary analysis will test gene-level associations with individual phenotypes using the optimal sequence kernel association test (SKAT-O) framed into SAIGE-GENE+. The SKAT-O test leverages the advantages of burden tests and SKAT through a linear combination of their test statistics, the relative contribution of which are estimated by a correlation term [56]. We chose this method to balance the need to maximise power for genes that have a higher proportion of causal variants satisfying the burden test assumption, while preserving power for genes that may have fewer causal variants (or variants with heterogeneous effects) despite our filtering strategy. The SAIGE-GENE+ method builds upon SKAT-O to reduce type I error inflation for very rare variants in large biobanks with unbalanced case-control ratios, reduce computational resources and account for sample relatedness [57]. We will use a relatedness coefficient cut-off of ≥0.125 (up to third-degree relatedness) in SAIGE-GENE+, and perform our analyses using the open-source R package *SAIGE* (https://github.com/saigegit/SAIGE). We will include the first 10 genetic principal components in the gene-level (combined with the generalized mixed model approach in SAIGE-GENE+) and RVGRS models to control for population structure [58], in addition to adjusting for age and sex [59]. We will also adjust for scanner site in neuroradiological analyses to control for potential technical confounding [60]. We will run separate gene-level tests for LOF/dominant negative and GOF variants to account for their anticipated opposite effect directions. As SKAT-O is designed to test the overall gene-trait association and does not produce effect sizes, variant-level effects will be obtained through separate logistic and linear regressions to help interpret p-values (as in Genebass). Genetic units with <10 carriers of any variant in the UKB will not be analysed to preserve power.

Our score and individual genes will be tested for their association with each phenotype of interest (n=18), and those with ≥1 statistically significant association with any phenotype of interest will be tested for associations across the phenome (n=196). We will interpret statistical significance in our score-based analysis with a Bonferroni-corrected p-value threshold to account for multiple testing across phenotypes (phenotypes of interest: 0.05/18=2.78x10^-3^; phenome: 0.05/196=2.55x10^-4^). We will interpret statistical significance in our gene-level analysis similarly, with a more stringent correction to account for multiple testing across genes and phenotypes (0.05/[# phenotypes x # gene-level units]) [61]. Our analyses will be conducted on the UK Biobank Research Analysis Platform [62]. We present our pre-planned sensitivity analyses in **Supplemental methods 2**.

### Power calculations

We performed a statistical power analysis for our gene-level tests and phenotypes of interest with SKAT-O using the *SKAT* package (v2.2.5) for R. Our results and analysis parameters are presented in **Figure 3** and detailed in **Supplemental methods 3**. Gene-level tests for lupus and vascular dementia have the lowest powers overall although they increase to reasonable values in more optimistic scenarios. Other cardiovascular and inflammatory outcomes have the highest power throughout all scenarios.

**Figure 3.**
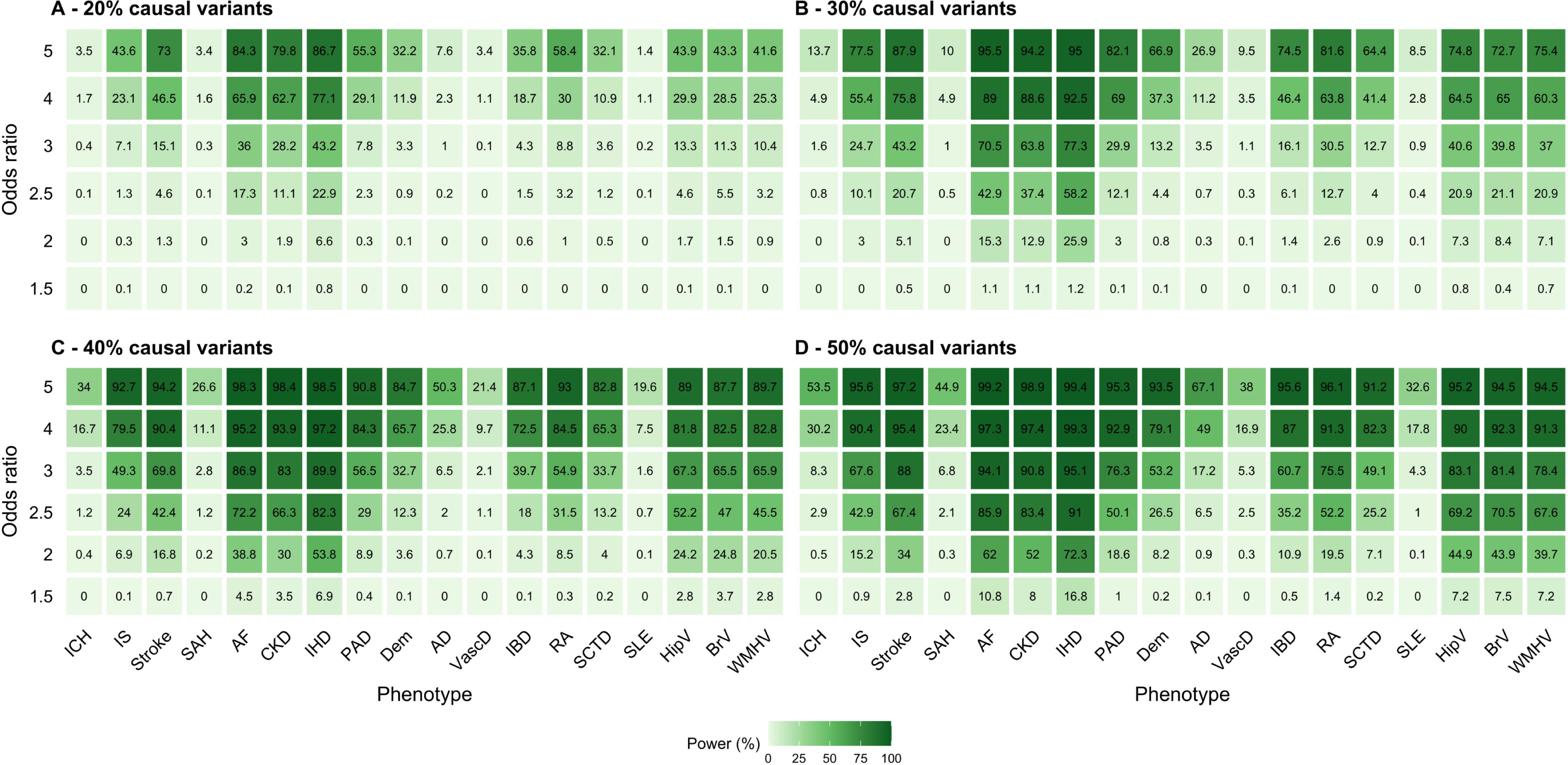
Power calculations for gene-level tests with phenotypes of interest using SKAT-O. The power calculation assumes an α=1.11x10^-5^, a genetic sampling length of 2,962 bp, a MAF <0.1%, an empirical optimal correlation coefficient, and sample sizes observed in the UKB. Abbreviations: AD, Alzheimer’s disease; AF, atrial fibrillation; bp, base pairs; BrV, total brain (grey plus white matter) volume; CKD, chronic kidney disease; Dem, all-cause dementia; HipV, hippocampal grey matter volume (average); IBD, inflammatory bowel disease; ICH, intracerebral haemorrhage; IHD, ischemic heart disease; IS, ischemic stroke; MAF, minor allele frequency; PAD, peripheral artery disease; RA, rheumatoid arthritis; SAH, subarachnoid haemorrhage; SCTD, systemic connective tissue disorder; SLE, systemic lupus erythematosus; VascD, vascular dementia; WMHV, total white matter hyperintensity volume.

## DISCUSSION

Comprehensive phenotyping of interferonopathy variant carriers may expand the clinical spectrum of genetic type I interferonopathies and help understand the biological relevance of type I interferon dysregulation in the general population. Importantly, large population-based assessments of interferonopathy carriers are lacking. Our study will leverage knowledge of Mendelian diseases of type I interferon to develop an informed, hypothesis-driven candidate pathway approach to investigate the frequency and phenotype associations of low-grade type I interferon dysregulation in the UKB. Our results will help understand the clinical spectrum of genetic type I interferonopathies, and will provide insights into the role of type I interferon in sporadic conditions.

Recent meta-analyses of genome-wide association studies (GWASs) have strengthened the case of inflammatory contributors to stroke and dementia. The largest cross-ancestry GWAS meta-analysis on stroke to date identified 89 independent genomic risk loci, of which two newly reported loci were located near or within genes involved in type I interferon regulation or signalling (*PTPN11* and *TAP1*) [63]. A recent large GWAS meta-analysis on Alzheimer’s disease and related dementias identified 33 known and 42 new genomic loci, for which a pathway analysis exposed significant gene sets related to immunity, including macrophage and microglia activation [64]. The nearest genes of two new lead variants, *SHARPIN* and *RBCK1*, encode essential components of the linear ubiquitin chain assembly complex (LUBAC), involved in NF-κB activation. Despite these discoveries, GWASs are unable to identify rarer alleles that may carry important information on the biology of complex traits, while most variants in genomic risk loci are mapped outside coding regions and have unknown regulatory functions [65, 66].

### Strengths and limitations

The UKB is the largest whole-exome sequencing project to date, markedly improving power to detect associations from a limited number of rare, functional variants [67]. Our informed approach will leverage current knowledge on type I interferon biology to reduce noise and test biologically plausible hypotheses. This contrasts with prior hypothesis-free phenome-wide association studies using rare variants in the UKB such as Genebass [42] and PheWAS [68], which did not include clinical annotations, used uncurated phenotypes, introduced greater multiple-testing burden (∼4.5k and ∼17k phenotypes tested in Genebass and PheWAS, respectively), and often used small sample sizes (as few as 30 cases/phenotype in PheWAS). The UKB also enables phenotyping from multiple sources, improving the classification accuracy for stroke and dementia as compared to studies using minimal phenotyping (e.g., case definition from self-reported dementia in relatives) [64].

Our study, however, will have some limitations. First, we expect that our weighting strategy based on theoretical knowledge will introduce noise into our score. We were technically unable to reliably assign empirical weights because of the lack of relevant transcript or protein measurements in the UKB. We anticipate this noise will be reduced by carefully selecting genes for which variants have a higher likelihood of functional and clinical consequences. We will also test genes individually as an alternative that does not mandate weights. Second, we anticipate some degree of residual pleiotropy through overlapping inflammatory and non-inflammatory pathways despite our careful curation of genes to increase specificity to the type I interferon cascade. We will, however, explore the relevance of pleiotropic effects on our results with a proteomic sensitivity analysis (**Supplemental methods 2**). Third, although we will optimize our overall power by carefully selecting clinically relevant genes and functional variants, our power will likely remain lower for rarer phenotypes.

## ETHICS AND DISSEMINATION

The UKB has received ethical approval from the North West Multicentre Research Ethics Committee, and all participants provided written informed consent at recruitment. This research will be conducted using the UKB Resource under application number 93,160. We expect to disseminate our results in a peer-reviewed journal and at an international cardiovascular conference.

## Data Availability

All data produced in the present study are available upon reasonable request to the authors.

## AUTHORS’ CONTRIBUTIONS

BR planned the study, performed the analyses, and drafted the protocol. WW and DH had a major role in the conception of the study, provided methodological and content expertise, and revised the manuscript. MC, RW, SM, KR, DM, JM, RB and YC provided methodological and content expertise, and had a major role in revising the manuscript. BR is the guarantor of the content of the study.

