## Supplemental for "Phenotypes associated with genetic determinants of type I interferon regulation in the UK Biobank: a protocol"

### Title

### Authors and affiliations

Bastien Rioux[1], Michael Chong[2,3,4], Rosie Walker[5], Sarah McGlasson[1], Kristiina Rannikmäe[6], Daniel McCartney[7], John McCabe[8,9], Robin Brown[10], Yanick J Crow[11,12], David Hunt[1*], William Whiteley[1,13*]

[1] Centre for Clinical Brain Sciences, University of Edinburgh, Edinburgh, United Kingdom

[2] Population Health Research Institute, McMaster University, Hamilton, Ontario, Canada

[3] Thrombosis and Atherosclerosis Research Institute, McMaster University, Hamilton, Ontario, Canada

[4] Department of Pathology and Molecular Medicine, McMaster University, Hamilton, Ontario, Canada

[5] Department of Psychology, University of Exeter, Exeter, United Kingdom

[6] Centre for Medical Informatics, Usher Institute, University of Edinburgh, Edinburgh, United Kingdom

[7] Centre for Genomic and Experimental Medicine, Institute of Genetics and Cancer, University of Edinburgh, Edinburgh, United Kingdom

[8] School of Medicine, University College Dublin, Dublin, Ireland

[9] Department of Medicine for the Elderly, Mater Misericordiae University Hospital, Dublin, Ireland

[10] Department of Clinical Neurosciences, University of Cambridge, Cambridge, United Kingdom

[11] MRC Human Genetics Unit, Institute of Genetics and Cancer, University of Edinburgh, Edinburgh, United Kingdom

[12] Laboratory of Neurogenetics and Neuroinflammation, Institut Imagine, Université de Paris, Paris, France

[13] MRC Population Health Unit, Nuffield Department of Population Health, University of Oxford

[*] Last authors with equal contribution

### SUPPLEMENTAL TABLES

**Supplemental table 1. Genes linked to type I interferonopathies (n=38).**

| Gene | Mutation effect (inheritance) | Main phenotype(s) | Proposed pathophysiology |
|---|---|---|---|
| *ACP5* | LOF (AR) | Spondyloenchondrodysplasia; spondyloenchondrodysplasia with immune dysregulation | Impaired TRAP enzyme fails to inactivate osteopontin, which consequently drives immune cells and osteoclasts |
| *ADAR* | LOF (AR) or DN (AD) | Aicardi-Goutières syndrome 6 | Impaired RNA editing enzyme that normally acts as a suppressor of type I IFN signalling |
| *ATAD3A* | LOF (AR) or DN (AD) | Global developmental delay, systemic sclerosis, spastic paraparesis | Cytosolic release of mitochondrial DNA |
| *ATM* | LOF (AR) | Ataxia telangiectasia | Impaired response to double-stranded DNA breaks and accumulation of DNA damage products |
| *BLM* | LOF (AR) | Bloom syndrome | Impaired RecQ–like helicase involved in genome stability resulting in the accumulation of DNA damage products |
| *C1QA* | LOF (AR) | Systemic lupus erythematosus | Unclear; involving $CD8^+$ T cell metabolism and downregulation of type I IFN production in plasmacytoid dendritic cells |
| *C1QB* | LOF (AR) | Systemic lupus erythematosus | |
| *C1QC* | LOF (AR) | Systemic lupus erythematosus | |
| *COPA* | DN (AD) | COPA syndrome | Impaired trafficking of STING from Golgi apparatus to endoplasmic reticulum |
| *DCLRE1C* | LOF (AR) | Immunodeficiency | Impaired DNA double-strand break repair resulting in the accumulation of DNA damage products |
| *DDX58* | GOF (AD) | Singleton-Merten syndrome 2 | Altered dsRNA recognition conferring a constitutive activation |
| *DNASE2* | LOF (AR) | Multisystem autoinflammatory syndrome with white matter hyperintensities, neonatal anaemia, glomerulonephritis, liver fibrosis, and deforming arthropathy | Reduced deoxyribonuclease activity results in impaired clearance of endogenous cytosolic nucleic acids |

| Gene | Mutation effect (inheritance) | Main phenotype(s) | Proposed pathophysiology |
|---|---|---|---|
| *IFIH1* | GOF (AD) | Aicardi-Goutières syndrome 7; Singleton-Merten syndrome 1 | Increased stability of the IFIH1 filament resulting in higher IFN signalling |
| *ISG15* | LOF (AR) | Immunodeficiency 38 with basal ganglia calcification (ISG15 deficiency) | Decreased function of ISG15, which normally prevents the degradation of an important negative regulator of IFN receptor signalling (USP18) |
| *JAK1* | GOF (AD) | Atopy, eosinophilia | Upregulated interferon-stimulated gene signalling |
| *LSM11* | LOF (AR) | Aicardi–Goutières syndrome 8 | Altered replication-dependent histone pre-mRNA processing leading to IFN stimulation by DNA via cGAS and STING |
| *NGLY1* | LOF (AR) | Early-onset encephalopathy | Impaired cellular clearance of damaged mitochondria |
| *PNPT1* | LOF (AR) | Early-onset encephalopathy, white matter hyperintensities, striatal necrosis | Altered import of RNA to mitochondria |
| *PSMA3* | LOF (AR) | Proteasome-associated autoinflammatory syndrome 1 | Impaired proteasome function leads to cell stress (namely through accumulation of cytosolic ubiquitinated proteins) |
| *PSMB4* | LOF (AR) | Proteasome-associated autoinflammatory syndrome 3 | |
| *PSMB8* | LOF (AR) | Proteasome-associated autoinflammatory syndrome 1 | |
| *PSMB9* | LOF (AR) | Proteasome-associated autoinflammatory syndrome 3 | |
| *PSMB10* | LOF (AR) | Proteasome-associated autoinflammatory syndrome 5 | |
| *PSMD12* | LOF (AD by haploinsufficiency) | Global developmental delay | |
| *PSMG2* | LOF (AR) | Proteasome-associated autoinflammatory syndrome 4 | |
| *POLA1* | LOF (X-linked recessive) | X-linked reticulate pigmentary disorder | Impaired DNA polymerase activity with subsequent change in cytosolic RNA-DNA hybrids |
| *POMP* | DN (AD) | Proteasome-associated autoinflammatory syndrome 2 | Impaired proteasome function leads to cell stress (namely through abnormal accumulation of ubiquitinated proteins) |

| Gene | Mutation effect (inheritance) | Main phenotype(s) | Proposed pathophysiology |
|---|---|---|---|
| *RNASEH2A* | LOF (AR) | Aicardi-Goutières syndrome 4 | Reduced RNase H2 function with subsequent change in cytosolic RNA-DNA hybrids |
| *RNASEH2B* | LOF (AR) | Aicardi-Goutières syndrome 2 | |
| *RNASEH2C* | LOF (AR) | Aicardi-Goutières syndrome 3 | |
| *RNU7-1* | LOF (AR) | Aicardi-Goutières syndrome 9 | Altered replication-dependent histone pre-mRNA processing leading to IFN stimulation by DNA via cGAS and STING |
| *SAMHD1* | LOF (AR) | Aicardi-Goutières syndrome 5 | Reduced nucleic acid metabolism and activation of cytosolic nucleic acid sensing mechanisms |
| *SKIC2* | LOF (AR) | Trichohepatoenteric syndrome 2 | Altered cytosolic RNA exosome function |
| *STAT1* | GOF (AD) | Immunodeficiency, autoimmunity, intracranial calcification | Impaired dephosphorylation in the nucleus |
| *STAT2* | LOF (AR) | Pseudo-TORCH syndrome 3 (STAT2-associated type I interferonopathy) | Decreased function of STAT2, which normally supports the function of an important negative regulator of type I IFN signalling (USP18) |
| *STING1* | GOF (AD) | STING-associated vasculopathy, infantile-onset | Abnormal upregulation of STING signalling, which increases production of IFN-β |
| *TREX1* | LOF (AR) or DN (AD) | Aicardi-Goutières syndrome 1 (AD mutations also cause retinal vasculopathy with cerebral leukoencephalopathy and systemic manifestations or RVCL-S) | Reduced deoxyribonuclease activity results in impaired clearance of endogenous cytosolic nucleic acids |
| *USP18* | LOF (AR) | Pseudo-TORCH syndrome 2 (USP18 deficiency) | Alteration of an enzyme that acts as a negative regulator of type I IFN signalling |

Abbreviations: AD, autosomal dominant; AR, autosomal recessive; cGAS, cyclic GMP-AMP synthase; DN, dominant-negative; DNA, deoxyribonucleic acid; GOF, gain-of-function; IFN, interferon; LOF, loss-of-function; RNA, ribonucleic acid; TORCH, toxoplasmosis-others-rubella-cytomegalovirus-herpes simplex; STING, cGAS–stimulator of interferon genes. Adapted from: Crow YJ et al, 2015 [1]; Crow YJ et al, 2022 [2]; Eleftheriou D et al, 2017 [3]; Rodero MP et al, 2016 [4].

**Supplemental table 2. Gene Ontology terms used to identify genes of interest (n=31).**

| ID | Term |
|---|---|
| GO:0071357 | cellular response to type I interferon |
| GO:0019962 | type I interferon binding |
| GO:0005132 | type I interferon receptor binding |
| GO:0004905 | type I interferon receptor activity |
| GO:0038197 | type I interferon receptor complex |
| GO:0035458 | cellular response to interferon-beta |
| GO:0035457 | cellular response to interferon-alpha |
| GO:0035456 | response to interferon-beta |
| GO:0035455 | response to interferon-alpha |
| GO:0060340 | positive regulation of type I interferon-mediated signaling pathway |
| GO:0060339 | negative regulation of type I interferon-mediated signaling pathway |
| GO:0060338 | regulation of type I interferon-mediated signaling pathway |
| GO:0060337 | type I interferon-mediated signaling pathway |
| GO:0072647 | interferon-epsilon production |
| GO:0072645 | interferon-delta production |
| GO:0072649 | interferon-kappa production |
| GO:0072651 | interferon-tau production |
| GO:0072653 | interferon-omega production |
| GO:0032728 | positive regulation of interferon-beta production |
| GO:0032727 | positive regulation of interferon-alpha production |
| GO:0032647 | regulation of interferon-alpha production |
| GO:0032648 | regulation of interferon-beta production |
| GO:0032687 | negative regulation of interferon-alpha production |
| GO:0032688 | negative regulation of interferon-beta production |
| GO:0034340 | response to type I interferon |
| GO:0032481 | positive regulation of type I interferon production |
| GO:0032480 | negative regulation of type I interferon production |
| GO:0032479 | regulation of type I interferon production |
| GO:0032607 | interferon-alpha production |
| GO:0032608 | interferon-beta production |
| GO:0032606 | type I interferon production |

**Supplemental table 3. Clinical and radiological phenotypes of interest (n=18).**

| Phenotype | N* | Classification source (UKB fields): diagnostic codes | Data sources (decreasing order of contribution) |
|---|---|---|---|
| **Cerebrovascular diseases (n=4)** | | | |
| Intracerebral haemorrhage | ~2,430 (algorithm: 2,310; primary care: 120) | UKB adjudication algorithm (42010, 42011):<br>**ICD v9**: 431.X (intracerebral haemorrhage);<br>**ICD v10**: I61 (intracerebral haemorrhage), I61.0 (intracerebral haemorrhage in hemisphere, subcortical), I61.1 (intracerebral haemorrhage in hemisphere, cortical), I61.2 (intracerebral haemorrhage in hemisphere, unspecified), I61.3 (intracerebral haemorrhage in brainstem), I61.4 (intracerebral haemorrhage in cerebellum), I61.5 (intracerebral haemorrhage, intraventricular), I61.6 (intracerebral haemorrhage, multiple localized), I61.8 (other intracerebral haemorrhage), I61.9 (intracerebral haemorrhage, unspecified);<br>**Self-report**: 20002/1491 (brain haemorrhage).<br>UKB first occurrence (131362, 131363):<br>**ICD v10**: I61 (intracerebral haemorrhage). | Inpatient hospital, death record, self-report, primary care |
| Ischemic stroke | ~12,074 (algorithm: 10,698; primary care: 1,376) | UKB adjudication algorithm (42008, 42009):<br>**ICD v9**: 434.X (occlusion of cerebral arteries), 434.0 (cerebral thrombosis), 434.1 (cerebral embolism), 434.9 (cerebral artery occlusion, unspecified), 436.X (acute, but ill-defined, cerebrovascular disease);<br>**ICD v10**: I63 (cerebral infarction), I63.0 (cerebral infarction due to thrombosis of precerebral arteries), I63.1 (cerebral infarction due to embolism of precerebral arteries), I63.2 (cerebral infarction due to unspecified occlusion or stenosis of precerebral arteries), I63.3 (cerebral infarction due to thrombosis of cerebral arteries), I63.4 (cerebral infarction due to embolism of cerebral arteries), I63.5 (cerebral infarction due to unspecified occlusion or stenosis of cerebral arteries), I63.6 (cerebral infarction due to cerebral venous thrombosis, nonpyogenic), I63.8 (other cerebral infarction), I63.9 (cerebral infarction, unspecified), I64.X (stroke, not specified as haemorrhage or infarction);<br>**Self-report**: 20002/1583 (ischaemic stroke). | Inpatient hospital, primary care, death record, self-report |

| Phenotype | N* | Classification source (UKB fields): diagnostic codes | Data sources (decreasing order of contribution) |
|---|---|---|---|
| | | UKB first occurrence (131366-131369):<br>**ICD v10**: I63 (cerebral infarction), I64 (stroke, not specified as haemorrhage or infarction). | |
| Stroke | ~19,771 (algorithm: 18,175; primary care: 1,596) | UKB adjudication algorithm (42006, 42007):<br>**ICD v9 and v10**: all diagnostic codes for intracerebral haemorrhage, ischemic stroke and subarachnoid haemorrhage;<br>**Self-report**: 20002/1081 (stroke), 20002/1086 (subarachnoid haemorrhage), 20002/1491 (brain haemorrhage), 20002/1583 (ischemic stroke).<br>Note: self-reported data does not include history of transient ischemic attack.<br>UKB first occurrence (131360-131363, 131366-131369):<br>**ICD v10**: I60 (subarachnoid haemorrhage), I61 (intracerebral haemorrhage), I63 (cerebral infarction), I64 (stroke, not specified as haemorrhage or infarction). | Inpatient hospital, self-report, primary care, death record |
| Subarachnoid haemorrhage | ~1,994 (algorithm: 1,894; primary care: 100) | UKB adjudication algorithm (42012, 42013):<br>**ICD v.9**: 430.X (subarachnoid haemorrhage);<br>**ICD v.10**: I60 (subarachnoid haemorrhage), I60.0 (subarachnoid haemorrhage from carotid siphon and bifurcation), I60.1 (subarachnoid haemorrhage from middle cerebral artery), I60.2 (subarachnoid haemorrhage from anterior communicating artery), I60.3 (subarachnoid haemorrhage from posterior communicating artery), I60.4 (subarachnoid haemorrhage from basilar artery), I60.5 (subarachnoid haemorrhage from vertebral artery), I60.6 (subarachnoid haemorrhage from other intracranial arteries), I60.7 (subarachnoid haemorrhage from intracranial artery, unspecified), I60.8 (other subarachnoid haemorrhage), I60.9 (subarachnoid haemorrhage, unspecified);<br>**Self-report**: 20002/1086 (subarachnoid haemorrhage).<br>UKB first occurrence (131360, 131361):<br>**ICD v10**: I60 (subarachnoid haemorrhage). | Inpatient hospital, self-report, death record, primary care |
| **Cardiovascular diseases (n=4)** | | | |

| Phenotype | N* | Classification source (UKB fields): diagnostic codes | Data sources (decreasing order of contribution) |
|---|---|---|---|
| Atrial fibrillation | 38,048 | UKB first occurrence (131350, 131351):<br>**ICD v10**: I48 (atrial fibrillation and flutter). | Inpatient hospital, primary care, self-report, death record |
| Chronic kidney disease | I12: 2,276;<br>I13: 195;<br>N18: 27,546;<br>N19: 3,743. | UKB first occurrence (131290-131293, 132032-132035):<br>**ICD v10**: I12 (hypertensive renal disease), I13 (hypertensive heart and renal disease), N18 (chronic kidney disease), N19 (unspecified kidney failure). | Inpatient hospital, primary care, self-report, death record |
| Ischemic heart disease | I20: 36,899;<br>I21: 23,587;<br>I22: 960;<br>I23: 102;<br>I24: 4,912;<br>I25: 51,807. | UKB first occurrence (131296-131307):<br>**ICD v10**: I20 (angina pectoris), I21 (acute myocardial infarction), I22 (subsequent myocardial infarction), I23 (certain current complications following acute myocardial infarction), I24 (other acute ischaemic heart disease), I25 (chronic ischaemic heart disease). | Inpatient hospital, self-report, primary care, death record |
| Peripheral artery disease | I70: 3,856;<br>I73: 13,740. | UKB first occurrence (131380, 131381, 131386, 131387):<br>**ICD v10**: I70 (atherosclerosis), I73 (other peripheral vascular diseases). | Inpatient hospital, primary care, self-report, death record |
| **Dementia (n=3)** | | | |
| All-cause dementia | ~7,951 (algorithm: 7,896; primary care: 55) | UKB adjudication algorithm (42018, 42019):<br>**ICD v9**: 290.2 (senile dementia, depressed or paranoid type), 290.3 (senile dementia with acute confusional state), 290.4 (arteriosclerotic dementia), 291.2 (other alcoholic dementia), 294.1 (dementia in other conditions classified elsewhere), 331.0 (Alzheimer's disease), 331.1 (Pick's disease), 331.2 (senile degeneration of brain), 331.5 (Creutzfeldt-Jakob disease);<br>**ICD v10**: A81.0 (sporadic Creutzfeldt-Jakob disease), F00 (dementia in Alzheimer's disease), F00.0 (dementia in Alzheimer's disease with early onset), F00.1 (dementia in Alzheimer's disease with late onset), F00.2 (dementia in Alzheimer's disease, atypical or mixed type), F00.9 (dementia in Alzheimer's disease, unspecified), F01 (vascular dementia), F01.0 (vascular dementia of acute onset), F01.1 (multi-infarct dementia), F01.2 (subcortical vascular dementia), F01.3 (mixed cortical and sub-cortical vascular dementia), F01.8 (other vascular dementia), F01.9 (vascular | Inpatient hospital, death record, self-report, primary care |

| Phenotype | N* | Classification source (UKB fields): diagnostic codes | Data sources (decreasing order of contribution) |
|---|---|---|---|
| | | dementia, unspecified), F02 (dementia in other diseases classified elsewhere), F02.0 (dementia in Picks disease), F02.1 (dementia in Creutzfeldt-Jacob disease), F02.2 (dementia in Huntington's disease), F02.3 (dementia in Parkinson's disease), F02.4 (dementia in HIV disease), F02.8 (dementia in other specified diseases classified elsewhere), F03 (unspecified dementia), F05.1 (delirium superimposed on dementia), F10.6 (mental and behavioural disorders due to use of alcohol – amnesic syndrome), G30 (Alzheimer's disease), G30.0 (Alzheimer's disease with early onset), G30.1 (Alzheimer's disease with late onset), G30.8 (other Alzheimer's disease), G30.9 (Alzheimer's disease unspecified), G31.0 (circumscribed brain atrophy), G31.1 (senile degeneration of brain), G31.8 (other specified degenerative diseases of nervous system);<br>**Self-report**: 20002/1263 (dementia/Alzheimer's/cognitive impairment).<br>UKB first occurrence (130836-130839, 130842, 130843, 131036, 131037):<br>**ICD v10**: F00 (dementia in Alzheimer's disease), F01 (vascular dementia), F03 (unspecified dementia), G30 (Alzheimer's disease). | |
| Alzheimer's disease | ~3,366 (algorithm: 3,290; primary care: 76) | UKB adjudication algorithm (42020, 42021):<br>**ICD v9**: 331.0 (Alzheimer's disease);<br>**ICD v10**: F00 (dementia in Alzheimer's disease), F00.0 (dementia in Alzheimer's disease), F00.1 (dementia in Alzheimer's disease with late onset), F00.2 (dementia in Alzheimer's disease, atypical or mixed type), F00.9 (dementia in Alzheimer's disease, unspecified), G30 (Alzheimer's disease), G30.0 (Alzheimer's disease with early onset), G30.1 (Alzheimer's disease with late onset), G30.8 (Other Alzheimer's disease), G30.9 (Alzheimer's disease unspecified).<br>UKB first occurrence (130836, 130837, 131036, 131037):<br>**ICD v10**: F00 (dementia in Alzheimer's disease), G30 (Alzheimer's disease). | Inpatient hospital, death record, primary care |

| Phenotype | N* | Classification source (UKB fields): diagnostic codes | Data sources (decreasing order of contribution) |
|---|---|---|---|
| Vascular dementia | ~1,830 (algorithm: 1,746; primary care: 84) | UKB adjudication algorithm (42022, 42023):<br>**ICD v9**: 290.4 (arteriosclerotic dementia);<br>**ICD v10**: F01 (vascular dementia), F01.0 (vascular dementia of acute onset), F01.1 (multi-infarct dementia), F01.2 (subcortical vascular dementia), F01.3 (mixed cortical and sub-cortical vascular dementia), F01.8 (other vascular dementia), F01.9 (vascular dementia, unspecified), I67.3 (Binswanger's disease).<br>UKB first occurrence (130838, 130839):<br>**ICD v10**: F01 (vascular dementia). | Inpatient hospital, death record, primary care |
| **Inflammatory and autoimmune diseases (n=4)** | | | |
| Inflammatory bowel disease | K50: 3,358; K51: 6,459. | UKB first occurrence (131626-131629):<br>**ICD v10**: K50 (Crohn's disease), K51 (ulcerative colitis). | Inpatient hospital, self-report, primary care, death record |
| Rheumatoid arthritis | M05: 1,401; M06: 12,589. | UKB first occurrence (131848-131851):<br>**ICD v10**: M05 (seropositive rheumatoid arthritis), M06 (other rheumatoid arthritis). | Inpatient hospital, self-report, primary care, death record |
| Systemic connective tissue disorder | M33: 275; M34: 469; M35: 7,335. | UKB first occurrence (131896-131901):<br>**ICD v10**: M33 (dermatopolymyositis), M34 (systemic sclerosis), M35 (other systemic involvement of connective tissue; includes Sjögren syndrome, Behçet disease, polymyalgia rheumatica). | Inpatient hospital, self-report, primary care, death record |
| Lupus erythematosus | M32: 1,053; L93: 517. | UKB first occurrence (131894, 131895, 131828, 131829):<br>**ICD v10**: M32 (systemic lupus erythematosus), L93 (lupus erythematosus; includes chilblain lupus). | Inpatient hospital, self-report, primary care, death record |

| Phenotype | N* | Classification source (UKB fields): diagnostic codes | Data sources (decreasing order of contribution) |
|---|---|---|---|
| **Radiological phenotypes (n=3)** | | | |
| Hippocampal grey matter volume | 42,935 | Volume of grey matter in hippocampus, average of right and left measurements (25886, 25887), $mm^3$ | Participants with brain magnetic resonance imaging |
| Total brain volume | 42,940 | Volume of brain (grey and white matter) normalized for head size (25009), $mm^3$ | |
| White matter hyperintensity volume | 41,623 | Total volume of white matter hyperintensities (25781), $mm^3$ | |

*$N_{cases}$ for binary outcomes, $N_{total}$ for continuous outcomes. N extracted from UK Biobank showcase (https://biobank.ctsu.ox.ac.uk/) as of January 2023. This table presents the clinical and radiological phenotypes of interest selected based on their frequency in the general population and the UKB, the plausibility of their association with type I IFN upregulation, and from type I interferonopathy presentations (including Mendelian and sporadic diseases). Abbreviations: ICD, International Classification of Diseases; IFN, interferon; UKB, UK Biobank.

**Supplemental table 4. Clinical phenotypes included in the phenome assessment (n=196).**

| Diagnostic code (ICD v10 unless specified) | $N_{cases}$ | Definition (UKB first occurrence field unless specified) |
|---|---|---|
| **Chapter 1: Certain infectious and parasitic diseases (n=22)** | | |
| A00-03, A05, A20-A28, A32-A36, A39, A42-44, A46, A48-A58, A66, A67, A69-A71, A74, A75, A77, A78, B95, B96 | See definition (right column) | Other bacterial infections:<br>A00 (cholera): n=26;<br>A01 (typhoid and paratyphoid fevers): n=165;<br>A02 (other salmonella infections): n=796;<br>A03 (shigellosis): n=212;<br>A05 (other bacterial foodborne intoxications): n=403;<br>A20 (plague): n=8;<br>A21 (tularaemia): n=0;<br>A22 (anthrax): n=2;<br>A23 (brucellosis): n=33;<br>A24 (glanders and melioidosis): n=1;<br>A25 (rat-bite fevers): n=3;<br>A26 (erysipeloid): n=13;<br>A27 (leptospirosis): n=35;<br>A28 (other zoonotic bacterial diseases, not elsewhere classified): n=65;<br>A32 (listeriosis): n=27;<br>A33 (tetanus neonatorum): n=1;<br>A34 (obstetrical tetanus): n=0;<br>A35 (other tetanus): n=119;<br>A36 (diphtheria): n=153;<br>A39 (meningococcal infection): n=117;<br>A42 (actinomycosis): n=219;<br>A43 (nocardiosis): n=3;<br>A44 (bartonellosis): n=1;<br>A46 (erysipelas): n=328;<br>A48 (other bacterial diseases, not elsewhere classified): n=204;<br>A49 (bacterial infection of unspecified site): n=2,885;<br>A50 (congenital syphilis): n=9;<br>A51 (early syphilis): n=20; |

| Diagnostic code (ICD v10 unless specified) | $N_{cases}$ | Definition (UKB first occurrence field unless specified) |
|---|---|---|
| | | A52 (late syphilis): n=21;<br>A53 (other and unspecified syphilis): n=30;<br>A54 (gonococcal infection): n=282;<br>A55 (chlamydial lymphogranuloma (venereum)): n=4;<br>A56 (other sexually transmitted chlamydial diseases): n=84;<br>A57 (chancroid): n=0;<br>A58 (granuloma inguinale): n=2;<br>A66 (yaws): n=24;<br>A67 (pinta [carate]): n=6;<br>A69 (other spirochaetal infections): n=320;<br>A70 (chlamydia psittaci infection): n=32;<br>A71 (trachoma): n=35;<br>A74 (other diseases caused by chlamydiae): n=40;<br>A75 (typhus fever): n=5;<br>A77 (spotted fever [tick-borne rickettsioses]): n=21;<br>A78 (q fever): n=10;<br>A79 (other rickettsioses): n=2;<br>B95 (streptococcus and staphylococcus as the cause of diseases classified to other chapters): n=8,987;<br>B96 (other bacterial agents as the cause of diseases classified to other chapters): n=18,726. |
| A60, A80, A90-99, B03, B04, B07-B09, B25, B27, B30, B33, B34, B91, B97, G14 | See definition | Other viral infections:<br>A60 (anogenital herpesviral [herpes simplex] infections): n=446;<br>A80 (acute poliomyelitis): n=538;<br>A90 (dengue fever [classical dengue]): n=65;<br>A91 (dengue haemorrhagic fever): n=2;<br>A92 (other mosquito-borne viral fevers): n=14;<br>A93 (other arthropod-borne viral fevers, not elsewhere classified): n=9;<br>A94 (unspecified arthropod-borne viral fever): n=4;<br>A95 (yellow fever): n=76;<br>A96 (arenaviral haemorrhagic fever): n=0;<br>A97 (dengue): n=35;<br>A98 (other viral haemorrhagic fevers, not elsewhere classified): n=1;<br>A99 (unspecified viral haemorrhagic fever): n=0; |

| Diagnostic code (ICD v10 unless specified) | $N_{cases}$ | Definition (UKB first occurrence field unless specified) |
|---|---|---|
| | | B03 (smallpox): n=14;<br>B04 (monkeypox): n=0;<br>B07 (viral warts): n=17,823;<br>B08 (other viral infections characterised by skin and mucous membrane lesions, not elsewhere classified): n=1,048;<br>B09 (unspecified viral infection characterised by skin and mucous membrane lesions): n=30;<br>B25 (cytomegaloviral disease): n=300;<br>B27 (infectious mononucleosis): n=3,604;<br>B30 (viral conjunctivitis): n=208;<br>B33 (other viral diseases, not elsewhere classified): n=240;<br>B34 (viral infection of unspecified site): 12,151;<br>B91 (sequelae of poliomyelitis): n=243;<br>B97 (viral agents as the cause of diseases classified to other chapters): n=3,774;<br>G14 (postpolio syndrome): n=84. |
| A06, A07, A59, B50-B60, B64-B83, B85-B89 | See definition | <u>Other parasitic infections</u>:<br>A06 (amoebiasis): n=118;<br>A07 (other protozoal intestinal diseases): n=513;<br>A59 (trichomoniasis): n=90;<br>B50 (plasmodium falciparum malaria): n=194;<br>B51 (plasmodium vivax malaria): n=43;<br>B52 (plasmodium malariae malaria): n=5;<br>B53 (other parasitologically confirmed malaria): n=5;<br>B54 (unspecified malaria): n=925;<br>B55 (leishmaniasis): n=17;<br>B56 (african trypanosomiasis): n=0;<br>B57 (chagas' disease): n=1;<br>B58 (toxoplasmosis): n=63;<br>B59 (pneumocystosis): n=181;<br>B60 (other protozoal diseases, not elsewhere classified): n=14;<br>B64 (unspecified protozoal disease): n=0;<br>B65 (schistosomiasis [bilharziasis]): n=112;<br>B66 (other fluke infections): n=7;<br>B67 (echinococcosis): n=12; |

| Diagnostic code (ICD v10 unless specified) | $N_{cases}$ | Definition (UKB first occurrence field unless specified) |
|---|---|---|
| | | B68 (taeniasis): n=12;<br>B69 (cysticercosis): n=6;<br>B70 (diphyllobothriasis and sparganosis): n=0;<br>B71 (other cestode infections): n=14;<br>B72 (dracunculiasis): n=0;<br>B73 (onchocerciasis): n=17;<br>B74 (filariasis): n=4;<br>B75 (trichinellosis): n=1;<br>B76 (hookworm diseases): n=33;<br>B77 (ascariasis): n=12;<br>B78 (strongyloidiasis): n=8;<br>B79 (trichuriasis): n=5;<br>B80 (enterobiasis): n=774;<br>B81 (other intestinal helminthiases, not elsewhere classified): n=17;<br>B82 (unspecified intestinal parasitism): n=96;<br>B83 (other helminthiases): n=85;<br>B85 (pediculosis and phthiriasis): n=339;<br>B86 (scabies): n=2,458;<br>B87 (myiasis): n=9;<br>B88 (other infestations): n=187;<br>B89 (unspecified parasitic disease): n=4. |
| A04 | 8,421 | Other bacterial intestinal infections |
| A08 | 5,610 | Viral and other specified intestinal infections |
| A09 | 24,576 | Diarrhoea and gastro-enteritis of presumed infectious origin |
| A15-A19, A30, A31, B90, B92 | See definition | Tuberculosis and other mycobacterial infections:<br>A15 (respiratory tuberculosis, bacteriologically and histologically confirmed): n=2,734;<br>A16 (respiratory tuberculosis, not confirmed bacteriologically or histologically): n=671;<br>A17 (tuberculosis of nervous system): n=22;<br>A18 (tuberculosis of other organs): n=196;<br>A19 (miliary tuberculosis): n=28;<br>A30 (leprosy [hansen's disease]): n=3;<br>A31 (infection due to other mycobacteria): n=160;<br>B90 (sequelae of tuberculosis): n=57; |

| Diagnostic code (ICD v10 unless specified) | $N_{cases}$ | Definition (UKB first occurrence field unless specified) |
|---|---|---|
| | | B92 (sequelae of leprosy): n=0. |
| A37 | 2,808 | Whooping cough |
| A38 | 1,711 | Scarlet fever |
| A40, A41 | See definition | Septicaemia:<br>A40 (streptococcal septicaemia): n=815;<br>A41 (other septicaemia): n=15,925. |
| A81-A89, G00-G03 | See definition | Viral meningitis and encephalitis:<br>A81 (atypical virus infections of central nervous system): n=67;<br>A82 (rabies): n=18;<br>A83 (mosquito-borne viral encephalitis): n=33;<br>A84 (tick-borne viral encephalitis): n=8;<br>A85 (other viral encephalitis, not elsewhere classified): n=15;<br>A86 (unspecified viral encephalitis): n=136;<br>A87 (viral meningitis): n=511;<br>A88 (other viral infections of central nervous system, not elsewhere classified): n=27;<br>A89 (unspecified viral infection of central nervous system): n=38;<br>G00 (bacterial meningitis, not elsewhere classified): n=235;<br>G01 (meningitis in bacterial diseases classified elsewhere): n=63;<br>G02 (meningitis in other infectious and parasitic diseases classified elsewhere): n=99;<br>G03 (meningitis due to other and unspecified causes): n=2,427. |
| B00 | 4,982 | Herpesviral [herpes simplex] infections |
| B01, B02 | See definition | Varicella infections:<br>B01 (varicella [chickenpox]): n=18,892;<br>B02 (zoster [herpes zoster]): n=18,325. |
| B05 | 13,383 | Measles |
| B06 | 4,924 | Rubella |
| B15-B19 | See definition | Viral hepatitis:<br>B15 (acute hepatitis a): n=1,136;<br>B16 (acute hepatitis b): n=422;<br>B17 (other acute viral hepatitis): n=465;<br>B18 (chronic viral hepatitis): n=878;<br>B19 (unspecified viral hepatitis): n=1,353. |

| Diagnostic code (ICD v10 unless specified) | $N_{cases}$ | Definition (UKB first occurrence field unless specified) |
|---|---|---|
| B20-B24 | See definition | Human immunodeficiency virus infection:<br>B20 (human immunodeficiency virus [hiv] disease resulting in infectious and parasitic diseases): n=70;<br>B21 (human immunodeficiency virus [hiv] disease resulting in malignant neoplasms): n=16;<br>B22 (human immunodeficiency virus [hiv] disease resulting in other specified diseases): n=12;<br>B23 (human immunodeficiency virus [hiv] disease resulting in other conditions): n=40;<br>B24 (unspecified human immunodeficiency virus [hiv] disease): n=518. |
| B26 | 8,798 | Mumps |
| B35 | 33,070 | Dermatophytosis |
| B36 | 5,871 | Other superficial mycoses |
| B37 | 12,628 | Candidiasis |
| B38-49 | See definition | Other fungal infections:<br>B38 (coccidioidomycosis): n=4;<br>B39 (histoplasmosis): n=9;<br>B40 (blastomycosis): n=1;<br>B41 (paracoccidioidomycosis): n=0;<br>B42 (sporotrichosis): n=2;<br>B43 (chromomycosis and phaeomycotic abscess): n=4;<br>B44 (aspergillosis): n=462;<br>B45 (cryptococcosis): n=8;<br>B46 (zygomycosis): n=4;<br>B47 (mycetoma): n=25;<br>B48 (other mycoses, not elsewhere classified): n=30;<br>B49 (unspecified mycosis): n=533. |
| **Chapter 2: Neoplasms (n=19)** | | |
| C00-C25, C30-C34, C37-C39, C43-C58, C60-C75, C81-C86, C90-C95 | See definition | Any malignant neoplasm:<br>C00 (malignant neoplasm of lip): n=63;<br>C01 (malignant neoplasm of base of tongue): n=218;<br>C02 (malignant neoplasm of other and unspecified parts of tongue): n=326;<br>C03 (malignant neoplasm of gum): n=99;<br>C04 (malignant neoplasm of floor of mouth): n=80;<br>C05 (malignant neoplasm of palate): n=97; |

| Diagnostic code (ICD v10 unless specified) | $N_{cases}$ | Definition (UKB first occurrence field unless specified) |
|---|---|---|
| | | C06 (malignant neoplasm of other and unspecified parts of mouth): n=111;<br>C07 (malignant neoplasm of parotid gland): n=143;<br>C08 (malignant neoplasm of other and unspecified major salivary glands): n=41;<br>C09 (malignant neoplasm of tonsil): n=383;<br>C10 (malignant neoplasm of oropharynx): n=58;<br>C11 (malignant neoplasm of nasopharynx): n=53;<br>C12 (malignant neoplasm of pyriform sinus): n=50;<br>C13 (malignant neoplasm of hypopharynx): n=35;<br>C14 (malignant neoplasm of other and ill-defined sites in the lip, oral cavity and pharynx): n=28;<br>C15 (malignant neoplasm of oesophagus): n=1,320;<br>C16 (malignant neoplasm of stomach): n=969;<br>C17 (malignant neoplasm of small intestine): n=363;<br>C18 (malignant neoplasm of colon): n=5,879;<br>C19 (malignant neoplasm of rectosigmoid junction): n=627;<br>C20 (malignant neoplasm of rectum): n=2,563;<br>C21 (malignant neoplasm of anus and anal canal): n=311;<br>C22 (malignant neoplasm of liver and intrahepatic bile ducts): n=738;<br>C23 (malignant neoplasm of gallbladder): n=157;<br>C24 (malignant neoplasm of other and unspecified parts of biliary tract): n=240;<br>C25 (malignant neoplasm of pancreas): n=1,500;<br>C30 (malignant neoplasm of nasal cavity and middle ear): n=65;<br>C31 (malignant neoplasm of accessory sinuses): n=28;<br>C32 (malignant neoplasm of larynx): n=396;<br>C33 (malignant neoplasm of trachea): n=9;<br>C34 (malignant neoplasm of bronchus and lung): n=5,176;<br>C37 (malignant neoplasm of thymus): n=58;<br>C38 (malignant neoplasm of heart, mediastinum and pleura): n=24;<br>C39 (malignant neoplasm of other and ill-defined sites in the respiratory system and intrathoracic organs): n=4;<br>C43 (malignant melanoma of skin): n=5,367;<br>C44 (other malignant neoplasms of skin): n=38,260;<br>C45 (mesothelioma): n=479; |

| Diagnostic code (ICD v10 unless specified) | $N_{cases}$ | Definition (UKB first occurrence field unless specified) |
|---|---|---|
| | | C46 (Kaposi's sarcoma): n=22;<br>C47 (malignant neoplasm of peripheral nerves and autonomic nervous system): n=23;<br>C48 (malignant neoplasm of retroperitoneum and peritoneum): n=215;<br>C49 (malignant neoplasm of other connective and soft tissue): n=469;<br>C50 (malignant neoplasm of breast): n=20,223;<br>C51 (malignant neoplasm of vulva): n=228;<br>C52 (malignant neoplasm of vagina): n=44;<br>C53 (malignant neoplasm of cervix uteri): n==603;<br>C54 (malignant neoplasm of corpus uteri): n=2,629;<br>C55 (malignant neoplasm of uterus, part unspecified): n=63;<br>C56 (malignant neoplasm of ovary): n=1,771;<br>C57 (malignant neoplasm of other and unspecified female genital organs): n=169;<br>C58 (malignant neoplasm of placenta): n=13;<br>C60 (malignant neoplasm of penis): n=105;<br>C61 (malignant neoplasm of prostate): n=15,260;<br>C62 (malignant neoplasm of testis): n=619;<br>C63 (malignant neoplasm of other and unspecified male genital organs): n=24;<br>C64 (malignant neoplasm of kidney, except renal pelvis): n=2,185;<br>C65 (malignant neoplasm of renal pelvis): n=138;<br>C66 (malignant neoplasm of ureter): n=137;<br>C67 (malignant neoplasm of bladder): n=1,942;<br>C68 (malignant neoplasm of other and unspecified urinary organs): n=43;<br>C69 (malignant neoplasm of eye and adnexa): n=223;<br>C70 (malignant neoplasm of meninges): n=24;<br>C71 (malignant neoplasm of brain): n=1,006;<br>C72 (malignant neoplasm of spinal cord, cranial nerves and other parts of central nervous system): n=35;<br>C73 (malignant neoplasm of thyroid gland): n=898;<br>C74 (malignant neoplasm of adrenal gland): n=45;<br>C75 (malignant neoplasm of other endocrine glands and related structures): n=27;<br>C81 (Hodgkin's disease): n=436;<br>C82 (follicular [nodular] non-Hodgkin's lymphoma): n=828;<br>C83 (diffuse non-Hodgkin's lymphoma): n=1,668; |

| Diagnostic code (ICD v10 unless specified) | $N_{cases}$ | Definition (UKB first occurrence field unless specified) |
|---|---|---|
| | | C84 (peripheral and cutaneous T-cell lymphomas): n=285;<br>C85 (other and unspecified types of non-Hodgkin's lymphoma): n=804;<br>C86 (other specified types of T/NK-cell lymphoma): n=6;<br>C90 (multiple myeloma and malignant plasma cell neoplasms): n=1,225;<br>C91 (lymphoid leukaemia): n=1,162;<br>C92 (myeloid leukaemia): n=767;<br>C93 (monocytic leukaemia): n=20;<br>C94 (other leukaemias of specified cell type): n=15;<br>C95 (leukaemia of unspecified cell type): n=27. |
| C00-C14 | See definition | <u>Malignant neoplasms of lip, oral cavity and pharynx</u>:<br>C00 (malignant neoplasm of lip): n=63;<br>C01 (malignant neoplasm of base of tongue): n=218;<br>C02 (malignant neoplasm of other and unspecified parts of tongue): n=326;<br>C03 (malignant neoplasm of gum): n=99;<br>C04 (malignant neoplasm of floor of mouth): n=80;<br>C05 (malignant neoplasm of palate): n=97;<br>C06 (malignant neoplasm of other and unspecified parts of mouth): n=111;<br>C07 (malignant neoplasm of parotid gland): n=143;<br>C08 (malignant neoplasm of other and unspecified major salivary glands): n=41;<br>C09 (malignant neoplasm of tonsil): n=383;<br>C10 (malignant neoplasm of oropharynx): n=58;<br>C11 (malignant neoplasm of nasopharynx): n=53;<br>C12 (malignant neoplasm of pyriform sinus): n=50;<br>C13 (malignant neoplasm of hypopharynx): n=35;<br>C14 (malignant neoplasm of other and ill-defined sites in the lip, oral cavity and pharynx): n=28. |
| C15-C17 | See definition | <u>Malignant neoplasms of upper gastrointestinal tract</u>:<br>C15 (malignant neoplasm of oesophagus): n=1,320;<br>C16 (malignant neoplasm of stomach): n=969;<br>C17 (malignant neoplasm of small intestine): n=363. |
| C18-C21 | See definition | <u>Malignant neoplasms of colon, rectum and anus</u>:<br>C18 (malignant neoplasm of colon): n=5,879;<br>C19 (malignant neoplasm of rectosigmoid junction): n=627; |

| Diagnostic code (ICD v10 unless specified) | $N_{cases}$ | Definition (UKB first occurrence field unless specified) |
|---|---|---|
| | | C20 (malignant neoplasm of rectum): n=2,563;<br>C21 (malignant neoplasm of anus and anal canal): n=311. |
| C22-C24 | See definition | Malignant neoplasms of liver, hepatic bile ducts and gallbladder:<br>C22 (malignant neoplasm of liver and intrahepatic bile ducts): n=738;<br>C23 (malignant neoplasm of gallbladder): n=157;<br>C24 (malignant neoplasm of other and unspecified parts of biliary tract): n=240. |
| C25 | 1,500 | Malignant neoplasm of pancreas |
| C30-C34, C37-C39 | See definition | Malignant neoplasms of respiratory and intrathoracic organs:<br>C30 (malignant neoplasm of nasal cavity and middle ear): n=65;<br>C31 (malignant neoplasm of accessory sinuses): n=28;<br>C32 (malignant neoplasm of larynx): n=396;<br>C33 (malignant neoplasm of trachea): n=9;<br>C34 (malignant neoplasm of bronchus and lung): n=5,176;<br>C37 (malignant neoplasm of thymus): n=58;<br>C38 (malignant neoplasm of heart, mediastinum and pleura): n=24;<br>C39 (malignant neoplasm of other and ill-defined sites in the respiratory system and intrathoracic organs): n=4. |
| C43 | 5,367 | Malignant melanoma of skin |
| C44 | 38,260 | Other malignant neoplasms of skin |
| C45-C49 | See definition | Malignant neoplasms of mesothelial and soft tissue:<br>C45 (mesothelioma): n=479;<br>C46 (Kaposi's sarcoma): n=22;<br>C47 (malignant neoplasm of peripheral nerves and autonomic nervous system): n=23;<br>C48 (malignant neoplasm of retroperitoneum and peritoneum): n=215;<br>C49 (malignant neoplasm of other connective and soft tissue): n=469. |
| C50 | 20,223 | Malignant neoplasm of breast |
| C51-C58 | See definition | Malignant neoplasms of female genital organs:<br>C51 (malignant neoplasm of vulva): n=228;<br>C52 (malignant neoplasm of vagina): n=44;<br>C53 (malignant neoplasm of cervix uteri): n==603;<br>C54 (malignant neoplasm of corpus uteri): n=2,629;<br>C55 (malignant neoplasm of uterus, part unspecified): n=63;<br>C56 (malignant neoplasm of ovary): n=1,771; |

| Diagnostic code (ICD v10 unless specified) | $N_{cases}$ | Definition (UKB first occurrence field unless specified) |
|---|---|---|
| | | C57 (malignant neoplasm of other and unspecified female genital organs): n=169;<br>C58 (malignant neoplasm of placenta): n=13. |
| C60-C63 | See definition | Malignant neoplasms of male genital organs:<br>C60 (malignant neoplasm of penis): n=105;<br>C61 (malignant neoplasm of prostate): n=15,260;<br>C62 (malignant neoplasm of testis): n=619;<br>C63 (malignant neoplasm of other and unspecified male genital organs): n=24. |
| C64-C68 | See definition | Malignant neoplasms of urinary tract:<br>C64 (malignant neoplasm of kidney, except renal pelvis): n=2,185;<br>C65 (malignant neoplasm of renal pelvis): n=138;<br>C66 (malignant neoplasm of ureter): n=137;<br>C67 (malignant neoplasm of bladder): n=1,942;<br>C68 (malignant neoplasm of other and unspecified urinary organs): n=43. |
| C69-C72 | See definition | Malignant neoplasms of eye, brain and other parts of central nervous system:<br>C69 (malignant neoplasm of eye and adnexa): n=223;<br>C70 (malignant neoplasm of meninges): n=24;<br>C71 (malignant neoplasm of brain): n=1,006;<br>C72 (malignant neoplasm of spinal cord, cranial nerves and other parts of central nervous system): n=35. |
| C73-C75 | See definition | Malignant neoplasms of thyroid and other endocrine glands:<br>C73 (malignant neoplasm of thyroid gland): n=898;<br>C74 (malignant neoplasm of adrenal gland): n=45;<br>C75 (malignant neoplasm of other endocrine glands and related structures): n=27. |
| C81-C86 | See definition | Lymphomas:<br>C81 (Hodgkin's disease): n=436;<br>C82 (follicular [nodular] non-Hodgkin's lymphoma): n=828;<br>C83 (diffuse non-Hodgkin's lymphoma): n=1,668;<br>C84 (peripheral and cutaneous T-cell lymphomas): n=285;<br>C85 (other and unspecified types of non-Hodgkin's lymphoma): n=804;<br>C86 (other specified types of T/NK-cell lymphoma): n=6. |
| C90 | 1,225 | Multiple myeloma and malignant plasma cell neoplasms |
| C91-C95 | See definition | Leukaemias:<br>C91 (lymphoid leukaemia): n=1,162; |

| Diagnostic code (ICD v10 unless specified) | $N_{cases}$ | Definition (UKB first occurrence field unless specified) |
|---|---|---|
| | | C92 (myeloid leukaemia): n=767;<br>C93 (monocytic leukaemia): n=20;<br>C94 (other leukaemias of specified cell type): n=15;<br>C95 (leukaemia of unspecified cell type): n=27. |
| **Chapter 3: Diseases of the blood and blood-forming organs and certain disorders involving the immune mechanism (n=9)** | | |
| D50-D53 | See definition | <u>Nutritional anaemias</u>:<br>D50 (iron deficiency anaemia): n=30,552;<br>D51 (vitamin b12 deficiency anaemia): n=3,710;<br>D52 (folate deficiency anaemia): n=941;<br>D53 (other nutritional anaemias): n=275. |
| D59-D64 | See definition | <u>Other acquired anaemias</u>:<br>D59 (acquired haemolytic anaemia): n=367;<br>D60 (acquired pure red cell aplasia [erythroblastopenia]): n=21;<br>D61 (other aplastic anaemias): n=1,567;<br>D62 (acute posthaemorrhagic anaemia): n=454;<br>D63 (anaemia in chronic diseases classified elsewhere): n=1,790;<br>D64 (other anaemias): 32,739. |
| D65, D68, D69 | See definition | <u>Acquired coagulation disorder</u>:<br>D65 (disseminated intravascular coagulation [defibrination syndrome]): n=98;<br>D68 (other coagulation defects): n=2,635;<br>D69 (purpura and other haemorrhagic conditions): 5,737. |
| D70 | 7,540 | Agranulocytosis |
| D71, D72 | See definition | <u>White blood cell disorders</u>:<br>D71 (functional disorders of polymorphonuclear neutrophils): n=34;<br>D72 (other disorders of white blood cells): n=1,783. |
| D73 | 1,327 | Diseases of spleen |
| D75 | 2,913 | Other diseases of blood and blood-forming organs |
| D80-D84 | See definition | <u>Immunodeficiency</u>:<br>D80 (immunodeficiency with predominantly antibody defects): n=433;<br>D81 (combined immunodeficiencies): n=16;<br>D82 (immunodeficiency associated with other major defects): n=49;<br>D83 (common variable immunodeficiency): n=53;<br>D84 (other immunodeficiencies): n=202. |

| Diagnostic code (ICD v10 unless specified) | $N_{cases}$ | Definition (UKB first occurrence field unless specified) |
|---|---|---|
| D86 | 2,109 | Sarcoidosis |
| **Chapter 4: Endocrine, nutritional and metabolic diseases (n=13)** | | |
| E03 | 38,989 | Other hypothyroidism (includes hypothyroidism, unspecified) |
| E05 | 8,402 | Thyrotoxicosis |
| E06 | 1,435 | Thyroiditis |
| E07 | 2,783 | Other disorders of thyroid |
| E10 | 5,182 | Insulin-dependent diabetes mellitus |
| E11 | 43,901 | Non-insulin-dependent diabetes mellitus |
| E10-E14 | See definition | Diabetes mellitus:<br>E10 (insulin-dependent diabetes mellitus): n=5,182;<br>E11 (non-insulin-dependent diabetes mellitus): n=43,901;<br>E12 (malnutrition-related diabetes mellitus): n=7;<br>E13 (other specified diabetes mellitus): n=545;<br>E14 (unspecified diabetes mellitus): n=25,676. |
| E20, E21 | See definition | Parathyroid disorder:<br>E20 (hypoparathyroidism): n=257;<br>E21 (hyperparathyroidism and other disorders of parathyroid gland): n=2,688. |
| E22 | 1,181 | Hyperfunction of pituitary gland |
| E23 | 1,257 | Hypofunction and other disorders of pituitary gland |
| E24-E27 | See definition | Adrenal gland disorder:<br>E24 (Cushing's syndrome): n=235;<br>E25 (adrenogenital disorders): n=36;<br>E26 (hyperaldosteronism): n=198;<br>E27 (other disorders of adrenal gland): n=1,646. |
| E28 | 2,343 | Ovarian dysfunction |
| E66 | 48,847 | Obesity |
| **Chapter 5: Mental and behavioural disorders (n=9)** | | |
| F20, F22, F23, F25, F28, F29 | See definition | Schizophrenia and related disorders:<br>F20 (schizophrenia): n=1,384);<br>F22 (persistent delusional disorders): n=626;<br>F23 (acute and transient psychotic disorders): n=321;<br>F25 (schizoaffective disorders): n=359; |

| Diagnostic code (ICD v10 unless specified) | $N_{cases}$ | Definition (UKB first occurrence field unless specified) |
|---|---|---|
| | | F28 (other nonorganic psychotic disorders): n=15;<br>F29 (unspecified nonorganic psychosis): n=642. |
| F31 | 2,561 | Bipolar affective disorder |
| F32 | 60,253 | Depressive episode |
| F33, F34, F38, F39 | See definition | Other mood disorders:<br>F33 (recurrent depressive disorder): n=3,839;<br>F34 (persistent mood [affective] disorders): n=916;<br>F38 (other mood [affective] disorders): n=167;<br>F39 (unspecified mood [affective] disorder): n=990. |
| F40 | 4,179 | Phobic anxiety disorders |
| F41 | 38,521 | Other anxiety disorders |
| F42 | 916 | Obsessive-compulsive disorder |
| F43 | 14,794 | Reaction to severe stress, and adjustment disorders |
| F44, F45, F48 | See definition | Somatoform and dissociation disorders:<br>F44 (dissociative [conversion] disorders): n=380;<br>F45 (somatoform disorders): n=3,106;<br>F48 (other neurotic disorders): n=1,152. |
| **Chapter 6: Diseases of the nervous system (n=10)** | | |
| G04, G05 | See definition | Encephalitis, myelitis and encephalomyelitis:<br>G04 (encephalitis, myelitis and encephalomyelitis): n=867;<br>G05 (encephalitis, myelitis and encephalomyelitis in diseases classified elsewhere): n=175. |
| G12 | 746 | Spinal muscular atrophy and related syndromes (including amyotrophic lateral sclerosis) |
| ICD v9 and v10; self-report | 3,807 | UKB adjudication algorithm, Parkinson's disease (42032, 42033):<br>ICD v9: 3320 (paralysis agitans);<br>ICD v10: G20 (Parkinson's disease);<br>Self-report: 20002/1262 (Parkinson's disease).<br>UKB first occurrence (131022, 131023):<br>ICD v10: G20 (Parkinson's disease). |
| G24 | 1,267 | Dystonia |
| G35 | 2,522 | Multiple sclerosis |
| G09, G36, G37, H46 | See definition | Other inflammatory disorders of the central nervous system:<br>G09 (sequelae of inflammatory diseases of central nervous system): n=105; |

| Diagnostic code (ICD v10 unless specified) | $N_{cases}$ | Definition (UKB first occurrence field unless specified) |
|---|---|---|
| | | G36 (other acute disseminated demyelination): n=38;<br>G37 (other demyelinating diseases of central nervous system): n=661;<br>H46 (optic neuritis): n=551. |
| G40, G41 | See definition | Epilepsy:<br>G40 (epilepsy): n=8,587;<br>G41 (status epilepticus): n=372. |
| G43 | 26,588 | Migraine |
| G44 | 6,070 | Other headache syndromes |
| G47 | 20,010 | Sleep disorders |
| **Chapter 7: Diseases of the eye and adnexa (n=8)** | | |
| H00, H01 | See definition | Inflammation of the eyelid:<br>H00 (hordeolum and chalazion): n=12,888;<br>H01 (other inflammation of eyelid): n=6,007. |
| H10 | 24,285 | Conjunctivitis |
| H15 | 1,329 | Disorders of sclera |
| H16 | 1,768 | Keratitis |
| H25, H26 | See definition | Cataract:<br>H25 (senile cataract): n=31,243;<br>H26 (other cataract): n=48,396. |
| H33 | 7,552 | Retinal detachments and breaks |
| H34 | 2,712 | Retinal vascular occlusions |
| H40 | 19,414 | Glaucoma |
| **Chapter 8: Diseases of the ear and mastoid process (n=2)** | | |
| H65-67 | See definition | Otitis media:<br>H65 (nonsuppurative otitis media): n=6,058;<br>H66 (suppurative and unspecified otitis media): n=10,291;<br>H67 (otitis media in diseases classified elsewhere): n=12. |
| H90 | 8,577 | Conductive and sensorineural hearing loss |
| **Chapter 9: Diseases of the circulatory system (n=24)** | | |
| I00, I01 | See definition | Acute rheumatic fever:<br>I00 (rheumatic fever without mention of heart involvement): 1,642;<br>I01 (rheumatic fever with heart involvement): n=32. |

| Diagnostic code (ICD v10 unless specified) | $N_{cases}$ | Definition (UKB first occurrence field unless specified) |
|---|---|---|
| I02, I05-I09 | See definition | Chronic rheumatic heart diseases:<br>I02 (rheumatic chorea): n=25;<br>I05 (rheumatic mitral valve diseases): n=907;<br>I06 (rheumatic aortic valve diseases): n=114;<br>I07 (rheumatic tricuspid valve diseases): n=1,763;<br>I08 (multiple valve diseases): n=7,709;<br>I09 (other rheumatic heart diseases): n=129. |
| I10 | 195,409 | Essential hypertension |
| I12 | 2,242 | Hypertensive renal disease |
| I26, I80-I82 | See definition | Venous thromboembolism:<br>I26 (pulmonary embolism): n=12,174;<br>I80 (phlebitis and thrombophlebitis): n=19,116;<br>I81 (portal vein thrombosis): n=468;<br>I82 (other venous embolism and thrombosis): n=1,356. |
| I27 | 2,952 | Other pulmonary heart disease (including primary pulmonary hypertension) |
| I30-I32 | See definition | Acute pericarditis and other diseases of pericardium:<br>I30 (acute pericarditis): n=713;<br>I31 (other diseases of pericardium): n=3,532;<br>I32 (pericarditis in diseases classified elsewhere): n=15. |
| I33, I38-I41 | See definition | Endocarditis and myocarditis:<br>I33 (acute and subacute endocarditis): n=610;<br>I38 (endocarditis, valve unspecified): n=1,110;<br>I39 (endocarditis and heart valve disorders in diseases classified elsewhere): n=12;<br>I40 (acute myocarditis): n=100;<br>I41 (myocarditis in diseases classified elsewhere): n=11. |
| I34 | 7,572 | Nonrheumatic mitral valve disorders |
| I35 | 8,343 | Nonrheumatic aortic valve disorders |
| I36, I37 | See definition | Nonrheumatic tricuspid and pulmonary valve disorders:<br>I36 (nonrheumatic tricuspid valve disorders): n=576;<br>I37 (pulmonary valve disorders): n=799. |
| I42, I43 | See definition | Cardiomyopathy:<br>I42 (cardiomyopathy): n=3,573;<br>I43 (cardiomyopathy in diseases classified elsewhere): n=131. |

| Diagnostic code (ICD v10 unless specified) | $N_{cases}$ | Definition (UKB first occurrence field unless specified) |
|---|---|---|
| I44, I45 | See definition | Conduction disorders:<br>I44 (atrioventricular and left bundle-branch block): n=12,387;<br>I45 (other conduction disorders): n=7,669. |
| I47 | 8,168 | Paroxysmal tachycardia |
| I49 | 13,643 | Other cardiac arrhythmias |
| I50 | 18,342 | Heart failure |
| I71, I72 | See definition | Aneurysm:<br>I71 (aortic aneurysm and dissection): n=4,712;<br>I72 (other aneurysm): n=1,594. |
| I74 | 2,143 | Arterial embolism and thrombosis |
| I77 | 3,513 | Other disorders of arteries and arterioles |
| I78 | 3,030 | Diseases of capillaries |
| I95 | 19,829 | Hypotension |
| ICD v9 and v10; self-report | 27,928 | UKB adjudication algorithm, myocardial infarction (42000, 42001):<br>ICD v9: 410 (acute myocardial infarction), 410.0 (acute myocardial infarction of anterolateral wall), 410.1 (acute myocardial infarction of other anterior wall), 410.2 (acute myocardial infarction of inferolateral wall), 410.3 (acute myocardial infarction of inferoposterior wall), 410.4 (acute myocardial infarction of other inferior wall), 410.5 (acute myocardial infarction of other lateral wall), 410.6 (true posterior wall infarction), 410.7 (subendocardial infarction), 410.8 (acute myocardial infarction of other specified sites), 410.9 (acute myocardial infarction of unspecified site), 411.0 (postmyocardial infarction syndrome), 412.X (old myocardial infarction), 429.79 (Ill-defined descriptions and complications of heart disease - other);<br>ICD v10: I21 (acute myocardial infarction), I21.0 (acute transmural myocardial infarction of anterior wall), I21.1 (acute transmural myocardial infarction of inferior wall), I21.2 (acute transmural myocardial infarction of other sites), I21.3 (acute transmural myocardial infarction of unspecified site), I21.4 (acute subendocardial myocardial infarction), I21.9 (acute myocardial infarction, unspecified), I22 (subsequent myocardial infarction), I22.0 (subsequent myocardial infarction of anterior wall), I22.1 (subsequent myocardial infarction of inferior wall), I22.8 (subsequent myocardial infarction of other sites), I22.9 (subsequent myocardial infarction of unspecified site), I23 (certain current complications following acute myocardial infarction), I23.0 (haemopericardium as current complication following acute myocardial infarction), I23.1 (atrial septal defect as current complication following acute |

| Diagnostic code (ICD v10 unless specified) | $N_{cases}$ | Definition (UKB first occurrence field unless specified) |
|---|---|---|
| | | myocardial infarction), I23.2 (ventricular septal defect as current complication following acute myocardial infarction), I23.3 (rupture of cardiac wall without haemopericardium as current complication following acute myocardial infarction), I23.4 (rupture of chordae tendineae as current complication following acute myocardial infarction), I23.5 (rupture of papillary muscle as current complication following acute myocardial infarction), I23.6 (thrombosis of atrium, auricular appendage, and ventricle as current complications following acute myocardial infarction), I23.8 (other current complications following acute myocardial infarction), I24.1 (Dressler syndrome), I25.2 (old myocardial infarction);<br>Self-report: 20002/1075 (heart attack/myocardial infarction).<br><u>UKB first occurrence (131296-131303, 131306, 131307)</u>:<br>ICD v10: I20 (angina pectoris), I21 (acute myocardial infarction), I23 (certain current complications following acute myocardial infarction), I25 (chronic ischaemic heart disease). |
| ICD v9 and v10; Read v2 and v3 | 16,750 | Rannikmäe et al [5]: stroke-specific or broad cerebrovascular codes from any medical setting |
| ICD v9 and v10; Read v2 and v3; self-report | 19,449 | Rannikmäe et al [5]: any stroke-specific or broad cerebrovascular codes from any medical setting and self-reported events |
| **Chapter 10: Diseases of the respiratory system (n=11)** | | |
| J00-J06 | See definition | <u>Acute upper respiratory tract infection</u>:<br>J00 (acute nasopharyngitis [common cold]): n=5,559;<br>J01 (acute sinusitis): n=24,620;<br>J02 (acute pharyngitis): n=24,231;<br>J03 (acute tonsillitis): n=18,549;<br>J04 (acute laryngitis and tracheitis): n=4,634;<br>J06 (acute upper respiratory infections of multiple and unspecified sites): n=52,282. |
| J09-J11 | See definition | <u>Influenza</u>:<br>J09 (influenza due to certain identified influenza virus): n=27;<br>J10 (influenza due to identified influenza virus): n=1,673;<br>J11 (influenza, virus not identified): n=11,044. |
| J12-J18 | See definition | <u>Pneumonia</u>:<br>J12 (viral pneumonia, not elsewhere classified): n=2,777;<br>J13 (pneumonia due to streptococcus pneumoniae): n=1,067;<br>J14 (pneumonia due to haemophilus influenzae): n=249;<br>J15 (bacterial pneumonia, not elsewhere classified): n=1,546; |

| Diagnostic code (ICD v10 unless specified) | $N_{cases}$ | Definition (UKB first occurrence field unless specified) |
|---|---|---|
| | | J16 (pneumonia due to other infectious organisms, not elsewhere classified): n=95;<br>J17 (pneumonia in diseases classified elsewhere): n=375;<br>J18 (pneumonia, organism unspecified): 36,373. |
| J20, J22 | See definition | Acute lower respiratory tract infection:<br>J20 (acute bronchitis): n=5,499;<br>J22 (unspecified acute lower respiratory infection): n=64,744. |
| J30 | 50,572 | Vasomotor and allergic rhinitis |
| J31-J33 | See definition | Chronic rhinosinusitis:<br>J31 (chronic rhinitis, nasopharyngitis and pharyngitis): n=6,987;<br>J32 (chronic sinusitis): n=13,697;<br>J33 (nasal polyp): n=7,829. |
| J41-J44 | See definition | Chronic obstructive pulmonary disease:<br>J41 (simple and mucopurulent chronic bronchitis): n=128;<br>J42 (unspecified chronic bronchitis): n=758;<br>J43 (emphysema): n=5,290;<br>J44 (other chronic obstructive pulmonary disease): n=26,153. |
| J45, J46 | See definition | Asthma:<br>J45 (asthma): n=72,870;<br>J46 (status asthmaticus): n=536. |
| J47 | 6,530 | Bronchiectasis |
| J84 | 4,136 | Other interstitial pulmonary diseases |
| J93 | 3,533 | Pneumothorax |
| **Chapter 11: Diseases of the digestive system (n=21)** | | |
| K02 | 6,278 | Dental caries |
| K05 | 2,164 | Gingivitis and periodontal diseases |
| K12 | 4,500 | Stomatitis and related lesions |
| K14 | 4,005 | Diseases of tongue |
| K20 | 19,039 | Oesophagitis |
| K21 | 76,989 | Gastro-oesophageal reflux disease |
| K25-K28 | See definition | Gastrointestinal ulcer:<br>K25 (gastric ulcer): n=12,777;<br>K26 (duodenal ulcer): n=9,824; |

| Diagnostic code (ICD v10 unless specified) | $N_{cases}$ | Definition (UKB first occurrence field unless specified) |
|---|---|---|
| | | K27 (peptic ulcer, site unspecified): n=1,917;<br>K28 (gastrojejunal ulcer): n=137. |
| K29 | 62,008 | Gastritis and duodenitis |
| K30 | 20,686 | Functional dyspepsia |
| K35-K37 | See definition | Appendicitis:<br>K35 (acute appendicitis): n=5,280;<br>K36 (other appendicitis): n=195;<br>K37 (unspecified appendicitis): n=6,821. |
| K52 | 29,580 | Other non-infective gastro-enteritis and colitis |
| K55 | 3,264 | Vascular disorders of intestine |
| K56 | 10,253 | Paralytic ileus and intestinal obstruction without hernia |
| K57 | 68,135 | Diverticular disease of intestine |
| K58, K59 | See definition | Irritable bowel syndrome:<br>K58 (irritable bowel syndrome): n=35,127;<br>K59 (other functional intestinal disorders): n=36,162. |
| K65 | 3,707 | Peritonitis |
| K70 | 2,254 | Alcoholic liver disease |
| K75 | 3,178 | Other inflammatory liver diseases |
| K80 | 33,890 | Cholelithiasis |
| K81 | 6,509 | Cholecystitis |
| K85 | 4,688 | Acute pancreatitis |
| **Chapter 12: Diseases of the skin and subcutaneous tissue (n=15)** | | |
| L00-L08 | See definition | Local infections of skin:<br>L00 (staphylococcal scalded skin syndrome): n=10;<br>L01 (impetigo): n=1,495;<br>L02 (cutaneous abscess, furuncle and carbuncle): n=12,932;<br>L03 (cellulitis): n=33,104;<br>L04 (acute lymphadenitis): n=788;<br>L05 (pilonidal cyst): n=1,624;<br>L08 (other local infections of skin and subcutaneous tissue): n=15,691. |
| L10-L14 | See definition | Bullous disorders:<br>L10 (pemphigus): n=97; |

| Diagnostic code (ICD v10 unless specified) | $N_{cases}$ | Definition (UKB first occurrence field unless specified) |
|---|---|---|
| | | L11 (other acantholytic disorders): n=122;<br>L12 (pemphigoid): n=266;<br>L13 (other bullous disorders): n=1,028;<br>L14 (bullous disorders in diseases classified elsewhere): n=1. |
| L20 | 13,899 | Atopic dermatitis |
| L21 | 10,073 | Seborrhoeic dermatitis |
| L40 | 15,425 | Psoriasis |
| L42 | 1,651 | Pityriasis rosea |
| L43 | 2,648 | Lichen planus |
| L50 | 11,194 | Urticaria |
| L70 | 6,285 | Acne |
| L71 | 6,146 | Rosacea |
| L80 | 1,210 | Vitiligo |
| L82 | 31,464 | Seborrhoeic keratosis |
| L90 | 6,437 | Atrophic disorders of skin |
| L91 | 5,271 | Hypertrophic disorders of skin |
| L92 | 1,970 | Granulomatous disorders of skin and subcutaneous tissue |
| **Chapter 13: Diseases of the musculoskeletal system and connective tissue (n=11)** | | |
| M00-M03 | See definition | <u>Infectious and reactive arthropathies</u>:<br>M00 (pyogenic arthritis): n=960;<br>M01 (direct infections of joint in infectious and parasitic diseases classified elsewhere): n=38;<br>M02 (reactive arthropathies): n=279;<br>M03 (postinfective and reactive arthropathies in diseases classified elsewhere): n=13. |
| M07-M09, M13 | See definition | <u>Other inflammatory arthropathies</u>:<br>M07 (psoriatic and enteropathic arthropathies): n=1,503;<br>M08 (juvenile arthritis): n=104;<br>M09 (juvenile arthritis in diseases classified elsewhere): n=2;<br>M13 (other arthritis): n=43,128. |
| M10 | 19,890 | Gout |
| M11 | 1,201 | Other crystal arthropathies |
| M15-M19 | See definition | <u>Arthrosis</u>: |

| Diagnostic code (ICD v10 unless specified) | $N_{cases}$ | Definition (UKB first occurrence field unless specified) |
|---|---|---|
| | | M15 (polyarthrosis): n=15,072;<br>M16 (coxarthrosis [arthrosis of hip]): n=26,800;<br>M17 (gonarthrosis [arthrosis of knee]): n=45,454;<br>M18 (arthrosis of first carpometacarpal joint): n=2,854;<br>M19 (other arthrosis): n=87,914. |
| M30, M31, L95 | See definition | Vasculitis:<br>M30 (polyarteritis nodosa and related conditions): n=222;<br>M31 (other necrotizing vasculopathies): n=2,039;<br>L95 (vasculitis limited to skin, not elsewhere classified): n=353. |
| M41 | 4,397 | Scoliosis |
| M45, M46 | See definition | Inflammatory spondylopathies:<br>M45 (ankylosing spondylitis): n=2,127;<br>M46 (other inflammatory spondylopathies): n=4,995. |
| M65 | 20,380 | Synovitis and tenosynovitis |
| M80-M82 | See definition | Osteoporosis:<br>M80 (osteoporosis with pathological fracture): n=2,453;<br>M81 (osteoporosis without pathological fracture): n=24,758;<br>M82 (osteoporosis in diseases classified elsewhere): n=190. |
| M86 | 2,769 | Osteomyelitis |
| **Chapter 14: Diseases of the genitourinary system (n=12)** | | |
| N00, N01, N03, N05 | See definition | Nephritic syndrome:<br>N00 (acute nephritic syndrome): n=298;<br>N01 (rapidly progressive nephritic syndrome): n=20;<br>N03 (chronic nephritic syndrome): n=1,286;<br>N05 (unspecified nephritic syndrome): n=837. |
| N10-N12 | See definition | Tubulo-interstitial nephritis:<br>N10 (acute tubulo-interstitial nephritis): n=1,613;<br>N11 (chronic tubulo-interstitial nephritis): n=363;<br>N12 (tubulo-interstitial nephritis, not specified as acute or chronic): n=2,692. |
| N13 | 7,136 | Obstructive and reflux uropathy |
| N17 | 22,204 | Acute renal failure |
| N20-N23 | See definition | Urolithiasis: |

| Diagnostic code (ICD v10 unless specified) | $N_{cases}$ | Definition (UKB first occurrence field unless specified) |
|---|---|---|
| | | N20 (calculus of kidney and ureter): n=12,295;<br>N21 (calculus of lower urinary tract): n=1,798;<br>N22 (calculus of urinary tract in diseases classified elsewhere): n=1;<br>N23 (unspecified renal colic): n=5,690. |
| N30 | 18,864 | Cystitis |
| N40 | 34,020 | Hyperplasia of prostate |
| N41, N45, N49 | See definition | Inflammatory diseases of male genital organs:<br>N41 (inflammatory diseases of prostate): n=4,860;<br>N45 (orchitis and epididymitis): n=3,080;<br>N49 (inflammatory disorders of male genital organs, not elsewhere classified): n=533. |
| N61 | 2,047 | Inflammatory disorders of breast |
| N70-N74 | See definition | Inflammatory diseases of female pelvic organs:<br>N70 (salpingitis and oophoritis): n=1,155;<br>N71 (inflammatory disease of uterus, except cervix): n=566;<br>N72 (inflammatory disease of cervix uteri): n=2,133;<br>N73 (other female pelvic inflammatory diseases): n=5,322;<br>N74 (female pelvic inflammatory disorders in diseases classified elsewhere): n=25. |
| N76 | 5,116 | Other inflammation of vagina and vulva (including vaginitis) |
| N80 | 10,025 | Endometriosis |
| **Chapter 15: Pregnancy, childbirth and the puerperium (n=4)** | | |
| O03 | 5,816 | Spontaneous abortion |
| O42 | 1,418 | Premature rupture of membranes |
| O68 | 4,331 | Labour and delivery complicated by foetal stress [distress] |
| O72 | 2,078 | Postpartum haemorrhage |
| **Chapter 16: Certain conditions originating in the perinatal period (n=0)** | | |
| NA | NA | NA |
| **Chapter 17: Congenital malformations, deformations and chromosomal abnormalities (n=6)** | | |
| Q00-Q07 | See definition | Congenital malformations of the nervous system:<br>Q00 (anencephaly and similar malformations): n=4;<br>Q01 (encephalocele): n=13;<br>Q02 (microcephaly): n=2;<br>Q03 (congenital hydrocephalus): n=42; |

| Diagnostic code (ICD v10 unless specified) | $N_{cases}$ | Definition (UKB first occurrence field unless specified) |
|---|---|---|
| | | Q04 (other congenital malformations of brain): n=98;<br>Q05 (spina bifida): n=397;<br>Q06 (other congenital malformations of spinal cord): n=44;<br>Q07 (other congenital malformations of nervous system): n=160. |
| Q10-Q18 | See definition | Congenital malformations of eye, ear, face and neck:<br>Q10 (congenital malformations of eyelid, lachrymal apparatus and orbit): n=144;<br>Q11 (anophthalmos, microphthalmos and macrophthalmos): n=32;<br>Q12 (congenital lens malformations): n=318;<br>Q13 (congenital malformations of anterior segment of eye): n=65;<br>Q14 (congenital malformations of posterior segment of eye): n=309;<br>Q15 (other congenital malformations of eye): n=55;<br>Q16 (congenital malformations of ear causing impairment of hearing): n=70;<br>Q17 (other congenital malformations of ear): n=129;<br>Q18 (other congenital malformations of face and neck): n=269. |
| Q20-Q28 | See definition | Congenital malformations of the circulatory system:<br>Q20 (congenital malformations of cardiac chambers and connexions): n=35;<br>Q21 (congenital malformations of cardiac septa): n=1,322;<br>Q22 (congenital malformations of pulmonary and tricuspid valves): n=64;<br>Q23 (congenital malformations of aortic and mitral valves): n=723;<br>Q24 (other congenital malformations of heart): n=444;<br>Q25 (congenital malformations of great arteries): n=282;<br>Q26 (congenital malformations of great veins): n=44;<br>Q27 (other congenital malformations of peripheral vascular system): n=346;<br>Q28 (other congenital malformations of circulatory system): n=271. |
| Q30-Q45, Q50-Q56 | See definition | Other congenital malformations:<br>Q30 (congenital malformations of nose): n=101;<br>Q31 (congenital malformations of larynx): n=75;<br>Q32 (congenital malformations of trachea and bronchus): n=18;<br>Q33 (congenital malformations of lung): n=75;<br>Q34 (other congenital malformations of respiratory system): n=12;<br>Q35 (cleft palate): n=42;<br>Q36 (cleft lip): n=10;<br>Q37 (cleft palate with cleft lip): n=43; |

| Diagnostic code (ICD v10 unless specified) | $N_{cases}$ | Definition (UKB first occurrence field unless specified) |
|---|---|---|
| | | Q38 (other congenital malformations of tongue, mouth and pharynx): n=629;<br>Q39 (congenital malformations of oesophagus): n=280;<br>Q40 (other congenital malformations of upper alimentary tract): n=309;<br>Q41 (congenital absence, atresia and stenosis of small intestine): n=6;<br>Q42 (congenital absence, atresia and stenosis of large intestine): n=24;<br>Q43 (other congenital malformations of intestine): n=576;<br>Q44 (congenital malformations of gallbladder, bile ducts and liver): n=298;<br>Q45 (other congenital malformations of digestive system): n=85;<br>Q50 (congenital malformations of ovaries, fallopian tubes and broad ligaments): n=319;<br>Q51 (congenital malformations of uterus and cervix): n=359;<br>Q52 (other congenital malformations of female genitalia): n=218;<br>Q53 (undescended testicle): n=597;<br>Q54 (hypospadias): n=289;<br>Q55 (other congenital malformations of male genital organs): n=159;<br>Q56 (indeterminate sex and pseudohermaphroditism): n=2. |
| Q60-Q64 | See definitions | <u>Congenital malformations of the urinary system</u>:<br>Q60 (renal agenesis and other reduction defects of kidney): n=353;<br>Q61 (cystic kidney disease): n=1,040;<br>Q62 (congenital obstructive defects of renal pelvis and congenital malformations of ureter): n=173;<br>Q63 (other congenital malformations of kidney): n=696;<br>Q64 (other congenital malformations of urinary system): n=192. |
| Q65-Q79 | See definitions | <u>Congenital malformations of musculoskeletal system</u>:<br>Q65 (congenital deformities of hip): n=346;<br>Q66 (congenital deformities of feet): n=1,169;<br>Q67 (congenital musculoskeletal deformities of head, face, spine and chest): n=324;<br>Q68 (other congenital musculoskeletal deformities): n=113;<br>Q69 (polydactyly): n=13;<br>Q70 (syndactyly): n=49;<br>Q71 (reduction defects of upper limb): n=31;<br>Q72 (reduction defects of lower limb): n=50;<br>Q73 (reduction defects of unspecified limb): n=10;<br>Q74 (other congenital malformations of limb(s)): n=283; |

| Diagnostic code (ICD v10 unless specified) | $N_{cases}$ | Definition (UKB first occurrence field unless specified) |
|---|---|---|
| | | Q75 (other congenital malformations of skull and face bones): n=42;<br>Q76 (congenital malformations of spine and bony thorax): n=841;<br>Q77 (osteochondrodysplasia with defects of growth of tubular bones and spine): n=41;<br>Q78 (other osteochondrodysplasias): n=212;<br>Q79 (congenital malformations of musculoskeletal system, not elsewhere classified): n=319. |

Phenotypes included are those available as first occurrences in the UK Biobank health-related outcomes (excluding cancer) and from cancer registries, unless specified. Underlined definitions group two or more diagnoses based on their pathophysiology and available cases. N extracted from UK Biobank showcase (https://biobank.ctsu.ox.ac.uk/) as of January 2023. Cancer numbers are for ICD v10 only and do not account for the small number of cancers coded with ICD v9. Abbreviations: ICD, International Classification of Diseases; UKB, UK Biobank.

### SUPPLEMENTAL METHODS

**Supplemental methods 1. Strengthening the Reporting of Genetic Association Studies (STREGA) checklist.**

| Item | # | STROBE guideline | Extension for genetic association studies (STREGA) | Manuscript section (page) |
|---|---|---|---|---|
| **Title and abstract** | 1 | (a) Indicate the study's design with a commonly used term in the title or the abstract. | | Abstract (p.3) |
| | | (b) Provide in the abstract an informative and balanced summary of what was done and what was found. | | NA |
| ***Introduction*** | | | | |
| **Background rationale** | 2 | Explain the scientific background and rationale for the investigation being reported. | | Introduction (p.5) |
| **Objectives** | 3 | State specific objectives, including any pre-specified hypotheses. | State if the study is the first report of a genetic association, a replication effort, or both. | Introduction (p.6) |
| ***Methods*** | | | | |
| **Study design** | 4 | Present key elements of study design early in the paper. | | Methods (p.6) |
| **Setting** | 5 | Describe the setting, locations and relevant dates, including periods of recruitment, exposure, follow-up, and data collection. | | Methods (p.6) |
| **Participants** | 6 | (a) *Cohort study*—Give the eligibility criteria, and the sources and methods of selection of participants. Describe methods of follow-up. *Case-control study*—Give the eligibility criteria, and the sources and methods of case ascertainment and control selection. Give the rationale for the choice of cases and controls. *Cross-sectional study*—Give the eligibility | Give information on the criteria and methods for selection of subsets of participants from a larger study, when relevant. | Methods (p.6) |

| | | | | |
|---|---|---|---|---|
| | | criteria, and the sources and methods of selection of participants. | | |
| | | (b) *Cohort study*—For matched studies, give matching criteria and number of exposed and unexposed.<br>*Case-control study*—For matched studies, give matching criteria and the number of controls per case. | | NA |
| **Variables** | 7 | (a) Clearly define all outcomes, exposures, predictors, potential confounders, and effect modifiers. Give diagnostic criteria, if applicable. | (b) Clearly define genetic exposures (genetic variants) using a widely used nomenclature system. Identify variables likely to be associated with population stratification (confounding by ethnic origin). | Methods (p.10-11, 15) |
| Data sources/ **measurement** | 8[a] | (a) For each variable of interest, give sources of data and details of methods of assessment (measurement). Describe comparability of assessment methods if there is more than one group. | (b) Describe laboratory methods, including source and storage of DNA, genotyping methods and platforms (including the allele calling algorithm used, and its version), error rates and call rates. State the laboratory/centre where genotyping was done. Describe comparability of laboratory methods if there is more than one group. Specify whether genotypes were assigned using all of the data from the study simultaneously or in smaller batches. | Methods (p.5-7) |
| **Bias** | 9 | (a) Describe any efforts to address potential sources of bias. | (b) For quantitative outcome variables, specify if any investigation of potential bias resulting from pharmacotherapy was undertaken. If relevant, describe the nature and magnitude of the potential bias, and explain what approach was used to deal with this. | NA |
| **Study size** | 10 | Explain how the study size was arrived at. | | Methods (p.16) |

| | | | | |
|---|---|---|---|---|
| **Quantitative variables** | 11 | Explain how quantitative variables were handled in the analyses. If applicable, describe which groupings were chosen, and why. | If applicable, describe how effects of treatment were dealt with. | Methods (p.13) |
| **Statistical methods** | 12 | (a) Describe all statistical methods, including those used to control for confounding. | State software version used and options (or settings) chosen. | Methods (p.14) |
| | | (b) Describe any methods used to examine subgroups and interactions. | | NA |
| | | (c) Explain how missing data were addressed. | | Methods (p.7-8) |
| | | (d) *Cohort study*—If applicable, explain how loss to follow-up was addressed.<br>*Case-control study*—If applicable, explain how matching of cases and controls was addressed.<br>*Cross-sectional study*—If applicable, describe analytical methods taking account of sampling strategy. | | Methods (p.8) |
| | | (e) Describe any sensitivity analyses. | | |
| | | | (f) State whether Hardy–Weinberg equilibrium was considered and, if so, how. | Methods (p.8) |
| | | | (g) Describe any methods used for inferring genotypes or haplotypes. | Methods (p.7) |
| | | | (h) Describe any methods used to assess or address population stratification. | Methods (p.15) |
| | | | (i) Describe any methods used to address multiple comparisons or to control risk of false positive findings. | Methods (p.15) |
| | | | (j) Describe any methods used to address and correct for relatedness among subjects. | Methods (p.14) |
| ***Results*** | | | | |
| **Participants** | 13[a] | (a) Report the numbers of individuals at each stage of the study—e.g., numbers potentially eligible, examined for eligibility, confirmed | Report numbers of individuals in whom genotyping was attempted and numbers | NA |

| | | | | |
|---|---|---|---|---|
| | | eligible, included in the study, completing follow-up, and analysed. | of individuals in whom genotyping was successful. | |
| | | (b) Give reasons for non-participation at each stage. | | NA |
| | | (c) Consider use of a flow diagram. | | NA |
| **Descriptive data** | 14[a] | (a) Give characteristics of study participants (e.g., demographic, clinical, social) and information on exposures and potential confounders. | Consider giving information by genotype. | NA |
| | | (b) Indicate the number of participants with missing data for each variable of interest. | | NA |
| | | (c) *Cohort study*—Summarize follow-up time, e.g. average and total amount. | | NA |
| **Outcome data** | 15[a] | *Cohort study*—Report numbers of outcome events or summary measures over time. | Report outcomes (phenotypes) for each genotype category over time. | NA |
| | | *Case-control study*—Report numbers in each exposure category, or summary measures of exposure. | Report numbers in each genotype category. | NA |
| | | *Cross-sectional study*—Report numbers of outcome events or summary measures. | Report outcomes (phenotypes) for each genotype category. | NA |
| **Main results** | 16 | (a) Give unadjusted estimates and, if applicable, confounder-adjusted estimates and their precision (e.g., 95% confidence intervals). Make clear which confounders were adjusted for and why they were included. | | NA |
| | | (b) Report category boundaries when continuous variables were categorized. | | NA |
| | | (c) If relevant, consider translating estimates of relative risk into absolute risk for a meaningful time period. | | NA |
| | | | (d) Report results of any adjustments for multiple comparisons. | NA |

| | | | | |
|---|---|---|---|---|
| **Other analyses** | 17 | (a) Report other analyses done—e.g., analyses of subgroups and interactions, and sensitivity analyses. | | NA |
| | | | (b) If numerous genetic exposures (genetic variants) were examined, summarize results from all analyses undertaken. | NA |
| | | | (c) If detailed results are available elsewhere, state how they can be accessed. | NA |
| ***Discussion*** | | | | |
| **Key results** | 18 | Summarize key results with reference to study objectives. | | NA |
| **Limitations** | 19 | Discuss limitations of the study, taking into account sources of potential bias or imprecision. Discuss both direction and magnitude of any potential bias. | | Discussion (p.17-18) |
| **Interpretation** | 20 | Give a cautious overall interpretation of results considering objectives, limitations, multiplicity of analyses, results from similar studies, and other relevant evidence. | | NA |
| **Generalizabilit y** | 21 | Discuss the generalizability (external validity) of the study results. | | NA |
| ***Other information*** | | | | |
| **Funding** | 22 | Give the source of funding and the role of the funders for the present study and, if applicable, for the original study on which the present article is based. | | Funding statement (p.31) |

This checklist only presents relevant sections for the protocol (other sections are assigned NA).

**Supplemental methods 2. Pre-planned sensitivity analyses.**

We will perform three sets of sensitivity analyses to help interpret our results. First, we will compare the risk of our 18 phenotypes of interest for groups of individuals:

i) in the highest versus lowest rare variant genetic risk score (RVGRS) decile;

ii) with a RVGRS raw value >0 versus a RVGRS raw value =0 and no variant;

iii) with a RVGRS raw value <0 versus a RVGRS raw value =0 and no variant.

This will help explore potential misclassifications of exposure in people with a null raw score, which may result from the absence of variants or the combination of variants with positive and negative weights. We will also re-run the RVGRS analysis after accounting for allelic phase and mode of inheritance, as our primary analysis assumes an additive architecture. The count value ($V_{i,e}$) of dominant and biallelic recessive variants (either homozygotes or trans-compound heterozygotes) will be increased by one to account for their stronger likelihood of pathogenicity. We will use a significance threshold of 0.05/(18 phenotypes x 4 tests)=$6.94 \times 10^{-4}$ to account for multiple testing.

Second, we will replicate our assessments of the RVGRS and genetic units with phenotypes of interest within disease-causing variants (i.e., excluding variants from functional annotations). This sensitivity analysis will help interpret potential noise introduced by variants included from in silico prediction. We will use the same significance threshold specified for primary and secondary analyses and interpret our results as exploratory.

Third, we will assess whether our results may be driven by a pleiotropic effect involving other canonical inflammatory pathways by testing the association of the RVGRS and genetic units with components of the inflammatory panel in Olink Explore 1536, a high-throughput protein biomarker discovery platform measuring 1,472 protein analytes used in ~54k UKB participants [6]. These include proteins for which upstream signalling may overlap with type I interferon [7], such as necrosis factor (TNF), interleukin 1α (IL-1 α), IL-1β, IL-6, IL-10, IL-18, IL-33, and C-reactive protein (CRP). We will also test the association of the RVGRS and genetic units with an interferon score derived from a sample of people with type I interferonopathies.

**Supplemental methods 3. Power analysis methodology.**

We performed a power analysis for our gene-level tests and phenotypes of interest (n=18) with SKAT-O using the *SKAT* package (v2.2.5) for R. We used an empirical optimal correlation coefficient (calculated from the observed proportion of causal variants), a minor allele frequency (MAF) <0.1%, and sample sizes observed in the UK Biobank for our phenotypes of interest. We anticipated our literature review to increase the number of genes from Gene Ontology (n=194) by ~25%, giving a total of ~250 included genes. We therefore set an a priori significance threshold $\alpha$=0.05/(18 phenotypes x 250 genes)=$1.11 \times 10^{-5}$. We used a genetic sampling length of 2,962 bp, corresponding to the median transcript length (i.e., exons plus untranslated regions) in the human genome, as we did not have a priori knowledge on transcript length in our gene set [8]. This variable is used to define the size of genetic regions randomly sampled 500 times across a default haplotype matrix (10k haplotypes over 200k bp) and to calculate an average power using the observed MAF. We calculated power across four proportions of causal variants (20-50%). Although this parameter is unknown beforehand, we expected a substantial enrichment in causal variants given our filtering strategy. We also assessed the impact of varying the maximum odds ratio (OR) observed (for a MAF=0.01%) from 5 (the default OR) down to a conservative OR of 1.5.
